# Seroprevalence of anti-SARS-CoV-2 antibodies and cross-variant neutralization capacity after the Omicron BA.2 wave in Geneva, Switzerland

**DOI:** 10.1101/2022.07.27.22278126

**Authors:** María-Eugenia Zaballa, Javier Perez-Saez, Carlos de Mestral, Nick Pullen, Julien Lamour, Priscilla Turelli, Charlène Raclot, Hélène Baysson, Francesco Pennacchio, Jennifer Villers, Julien Duc, Viviane Richard, Roxane Dumont, Claire Semaani, Andrea Jutta Loizeau, Clément Graindorge, Elsa Lorthe, Jean-François Balavoine, Didier Pittet, Manuel Schibler, Nicolas Vuilleumier, François Chappuis, Omar Kherad, Andrew S. Azman, Klara M. Posfay-Barbe, Laurent Kaiser, Didier Trono, Silvia Stringhini, Idris Guessous, the Specchio-COVID19 study group

## Abstract

**Background:** More than two years into the COVID-19 pandemic, it is generally assumed that most of the population has developed anti-SARS-CoV-2 antibodies from infection and/or vaccination. However, public health decision-making is hindered by the lack of up-to-date and precise characterization of the immune landscape in the population. We thus aimed to estimate anti-SARS-CoV-2 antibodies seroprevalence and cross-variant neutralization capacity after Omicron became dominant in Geneva, Switzerland.

**Methods:** We conducted a population-based serosurvey between April 29^th^ and June 9^th^, 2022, recruiting children and adults of all ages from age-stratified random samples of the Geneva general population. Anti-SARS-CoV-2 antibody presence was assessed using commercial immunoassays targeting either the spike (S) or nucleocapsid (N) protein. Antibodies neutralization capacity against different SARS-CoV-2 variants was evaluated using a cell-free Spike trimer-ACE2 binding-based surrogate neutralization assay. Seroprevalence of anti-SARS-CoV-2 antibodies and neutralization capacity were estimated using Bayesian modeling frameworks accounting for the demographics, vaccination, and infection statuses of the Geneva population.

**Results:** Among the 2521 individuals included in the analysis (55.2% women; 21.4% aged <18 years and 14.2% aged ≥ 65 years), overall seroprevalence of antibodies was 93.8% (95% credible interval: 93.1-94.5), including 72.4% (70.0-74.7) for infection-induced antibodies. Estimates of neutralizing antibodies based on a representative subsample of 1160 participants ranged from 79.5% (77.1-81.8) against the Alpha variant to 46.7% (43.0-50.4) against the Omicron BA.4/BA.5 subvariants. Despite having high seroprevalence of infection-induced antibodies (76.7% [69.7-83.0] for ages 0-5 years, 90.5% [86.5-94.1] for ages 6-11 years), children aged <12 years had substantially lower neutralizing activity than older participants, particularly against Omicron subvariants. In general, higher levels of neutralization activity against pre-Omicron variants were associated with vaccination, particularly having received a booster dose. Higher levels of neutralization activity against Omicron subvariants were associated with booster vaccination alongside recent infection.

**Conclusion:** More than nine in ten individuals in the Geneva population have developed anti-SARS-CoV-2 antibodies through vaccination and/or infection, but less than half of the population has antibodies with neutralizing activity against the currently circulating Omicron BA.5 subvariant. Hybrid immunity obtained through booster vaccination and infection appears to confer the greatest neutralization capacity, including against Omicron.

## INTRODUCTION

By the end of 2021, most of the world’s population had developed antibodies against the severe acute respiratory syndrome coronavirus 2 (SARS-CoV-2), through infection, vaccination, or both (1–3). In high-income countries, vaccination programs have contributed to high prevalence of anti-SARS-CoV-2 antibodies particularly among elderly people and individuals with chronic conditions (1–3), two groups with high risk of severe COVID-19, hospitalization, and death (4,5). While population-based serosurveys remain important to monitor the pandemic (6), they fall short in shedding light on the state of population-level protection against infection from current SARS-CoV-2 variants (7–10). In fact, studies on neutralizing antibodies, primarily based on small and non-representative samples (11), have shown that neutralizing capacity depends strongly on how antibodies are mounted (through vaccination and/or infection; and the number, duration and severity of infection episodes) and may differ between variants (8–11). This has implications for current immune landscapes shaped by complex age-specific vaccination and infection patterns. Vaccination-induced antibodies have demonstrated high neutralizing capacity against previously predominant variants (ancestral D614G, Alpha, and Delta), but substantially less neutralizing capacity against more recent and currently dominant Omicron subvariants (8,9,12,13)—even though, importantly, protection against severe disease, hospitalization, and death remains high (14,15). Simultaneously, antibodies developed through infection by the Omicron BA.1 subvariant have shown reduced neutralizing capacity against most other variants of concern (VOCs) (8,9,13,16,17). From a public health standpoint, having up-to-date estimates of antibody seroprevalence, their origin, and their neutralizing capacity against diverse circulating VOCs is critically important to disseminate public health messages to the population and to inform and adapt decisions on vaccination strategies, mask requirements, and other preventive measures. Such evidence-based decisions are needed to minimize the number of people falling ill with severe disease, requiring hospitalization, and dying during the current wave, driven by the highly contagious Omicron BA.5 subvariant, as well as future pandemic waves.

To our knowledge, there are no population-based seroprevalence estimates of neutralizing antibodies against main VOCs, particularly after the Omicron variant became predominant. To fill this gap, we recruited a representative sample of the general population of Geneva, Switzerland, and assessed seroprevalence of anti-SARS-CoV-2 antibodies and their neutralizing capacity against SARS-CoV-2 VOCs 28 months after the first confirmed case in the country and 5 months after the Omicron BA.1 subvariant became dominant (18).

With a population of about 500000, the canton of Geneva, Switzerland, has had 261946 confirmed cases (448 per 1000 inhabitants) and 883 deaths reported by July 21^st^, 2022 (19). Our previous serosurvey revealed that by June-July 2021, around two thirds of the population had developed anti-SARS-CoV-2 antibodies following vaccination and/or infection, half of which having antibodies of infection origin (20). Since then, eligibility for vaccination against COVID-19 has progressively expanded in Geneva to cover most age groups, including ages 12-15 years since June 2021 and ages 5-11 years since January 2022 (Supplementary figure 1).

## METHODS

### Study design

Between April 29^th^ and June 9^th^, 2022, we recruited participants from a random sample of individuals aged ≥ 6 months provided by the cantonal population registry (*Office cantonal de la population et des migrations*), and from an age- and sex-stratified random sample of adults who participated in at least one of our previous serosurveys (Supplementary figure 2) (20–22). Newly selected individuals were invited by letter, while returning participants were invited by letter or email when available. A written reminder was sent to all non-responding individuals and a phone call was made to all whose phone number was available. Children and teenagers (<18 years) were invited to participate with members of their household. Participation rates differed across age groups and depending on previous participation: 12.4% for those aged <18 years, 18.8% for those aged 18-64 years and 32.9% for those aged ≥ 65 years among newly invited participants; and 57.6% for those aged 18-64 years and 75.5% for those aged ≥ 65 years among returning participants (Supplementary figures 2-3). Participants provided a venous blood sample and completed an online or paper questionnaire that collected sociodemographic and vaccination information and COVID-19-related medical history. Informed written consent was obtained from all participants. The Geneva Cantonal Commission for Research Ethics approved this study (Project N° 2020-00881).

### Immunoassays

To detect anti-SARS-CoV-2 antibodies, we used two commercially-available immunoassays: the Roche Elecsys anti-SARS-CoV-2 S and anti-SARS-CoV-2 N immunoassays (Roche Diagnostics, Rotkreuz, Switzerland), which detect immunoglobulins (IgG/A/M) against the receptor binding domain of the virus spike (S) protein (#09 289 275 190, Roche-S) and the virus nucleocapsid (N) protein (#09 203 079 190, Roche-N), respectively. Both assays have high accuracy and have been validated in multiple settings, including our previous serosurveys (20–22). We defined seropositivity using the manufacturer’s provided cut-off values of titer ≥ 0.8 U/mL for the Roche-S, and cut-off index ≥ 1.0 for the Roche-N immunoassays.

### S^3^-cell free neutralization assay

To assess anti-SARS-CoV-2 antibody neutralizing activity, we used the S^3^-ACE2 neutralization assay (23,24). Production and purification of trimeric Spike variants (D614G [B.1], Alpha [B1.1.7], Beta [B.1.351], Gamma [P.1], Delta [B.1.617.2], Iota [B.1.526], Kappa [B.1.617.1], Lambda [C.37], and Omicron [BA.1, BA.2, BA.2.12.1 and BA.4/BA.5]) and ACE2 mouse Fc fusion protein as well as Spike protein-beads coupling were performed as previously described (23). Neutralization assays were done in 96-well plates, where 10 multiplexed Spike variants were incubated with sera, as described (23). Briefly, a volume of 5 μL of serum per well was used for the starting dilution and a total of 6 serial dilutions (1:10, 1:30, 1:90, 1:270, 1:810, 1:7290) of sera were incubated 1 hour with the Spike proteins before ACE2 mouse Fc fusion protein was added, and binding detected with an anti-mouse IgG-PE secondary antibody (eBioscience, Thermo Fisher Scientific, catalogue #12-4010-87). Control wells were included on each 96-well plate with each variant Spike-coupled beads alone. Plates were read with a Luminex 200 instrument and mean fluorescence intensity (MFI) for beads without serum was averaged and used as the 100% binding signal for the ACE2 receptor to the bead-coupled Spike trimer. MFI obtained for D614G Spike using a high concentration of imdevimab (RGN10987, 1 μg/mL), a monoclonal antibody known to neutralize the ancestral strain, was used as the maximum inhibition signal (25). The percent blocking of the Spike trimer-ACE2 interaction was calculated using the formula: % Inhibition = (1- ([MFI Test dilution – MFI Max inhibition] / [MFI Max binding – MFI Max inhibition]) × 100). Serum dilution response inhibition curves were generated using GraphPad Prism 8.3.0. NonLinear four-parameter curve fitting analysis of the agonist versus response, and ED50% values (mean serum dilution needed to achieve 50% neutralization) were extracted using an in-house script. Neutralizing capacity was assessed against the ancestral variant (D614G), the Alpha, Beta, Gamma, Delta, Iota, Kappa, and Lambda variants, and the Omicron BA.1, BA.2, BA.2.12.1, and BA.4/BA.5 subvariants (Spike proteins of BA.4 and BA.5 subvariants share identical sequences (17) and thus results of the cell-free surrogate neutralization assay apply to both).

### Statistical analyses

To estimate seroprevalence of anti-SARS-CoV-2 antibodies (% and 95% credible intervals [95% CrI]), we used a Bayesian modelling framework jointly inferring anti-N and anti-S presence while accounting for age, sex, immunoassay performance, and household clustering following our previous work (20). Since the vaccines used to date in Geneva do not elicit a response to the SARS-CoV-2 N-protein (26), we used participants’ two-marker antibody profiles to estimate the proportion of those having anti-SARS-CoV-2 antibodies from any origin (vaccination and/or infection) and those having antibodies due to infection (who could be vaccinated or not). We further developed a Bayesian logistic model to estimate the seroprevalence of neutralizing antibodies (% and 95% CrI), accounting for age, sex, and infection (uninfected, latest infected by a pre-Omicron variant, latest infected by an Omicron subvariant) and vaccination status (unvaccinated, vaccinated without booster, vaccinated with booster). Data on infection dates and vaccination was self-reported by the participants at the moment of the blood drawing (Supplementary information S1). Since Omicron became the dominant circulating variant in the Geneva region by late December 2021 (18), we assumed infections were due to Omicron (no subvariants distinction) if the participant reported having had a COVID-19 diagnostic positive test (RT-PCR or rapid antigen test, including self-test) after January 1^st^, 2022. Our base model assumed additive contributions of vaccination and infection and was fit to each variant separately. As sensitivity analysis, we fit three additional sets of models: the first with an interaction term between infection and vaccination status, the second with overdispersion in neutralizing capacity, and the third with differential surrogate neutralization test performance for children under the age of 12 years versus older individuals. Model selection was performed based on estimated leave-one-out cross-validation error (27). Population-level estimates of antibodies seroprevalence and neutralization capacity were obtained through post-stratification to the demographics and vaccination statuses of the Geneva general population. Full details of the statistical models are provided in Supplementary information S2.

## RESULTS

### Overall anti-SARS-CoV-2 seroprevalence estimates

Our analytical sample for seroprevalence estimation comprised 2521 participants (Supplementary Figure 2), of whom 55.2% were women, 21.4% were aged <18 years and 14.2% were aged ≥ 65 years (Supplementary figure 4). Among adults, 11.3% had a primary education level and 56.9% had a tertiary education level, compared with 26.6% and 41.5%, respectively, in the general population of Geneva (Supplementary table 1). Overall, 75.4% of participants declared having received at least one COVID-19 vaccine dose at the time of their recruitment in the study, compared with 71.1% in the Geneva general population (Supplementary table 2); 96.9% of all participants tested positive for anti-S antibodies, and 70.0% tested positive for anti-N antibodies (Table 1; Supplementary figure 5).

**Table 1.**
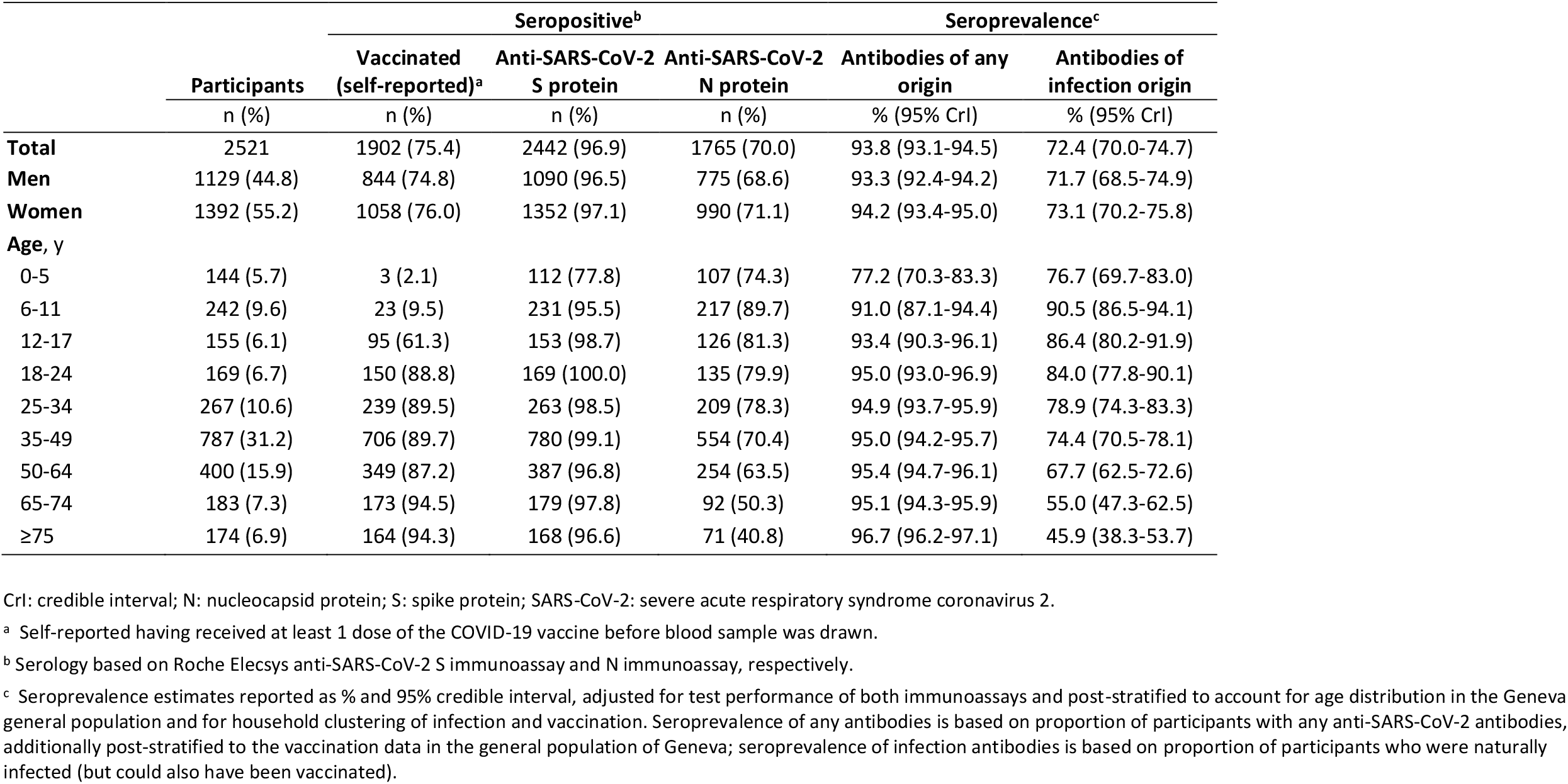
Demographic characteristics of sample, serological results, and seroprevalence estimates in Geneva, Switzerland, April 29^th^ to June 9^th^, 2022

|  | Participants | Seropositive<sup>b</sup> |  |  | Seroprevalence<sup>c</sup> |  |
| --- | --- | --- | --- | --- | --- | --- |
|  |  | Vaccinated (self-reported)<sup>a</sup> | Anti-SARS-CoV-2 S protein | Anti-SARS-CoV-2 N protein | Antibodies of any origin | Antibodies of infection origin |
|  | n (%) | n (%) | n (%) | n (%) | % (95% CrI) | % (95% CrI) |
| **Total** | 2521 | 1902 (75.4) | 2442 (96.9) | 1765 (70.0) | 93.8 (93.1-94.5) | 72.4 (70.0-74.7) |
| **Men** | 1129 (44.8) | 844 (74.8) | 1090 (96.5) | 775 (68.6) | 93.3 (92.4-94.2) | 71.7 (68.5-74.9) |
| **Women** | 1392 (55.2) | 1058 (76.0) | 1352 (97.1) | 990 (71.1) | 94.2 (93.4-95.0) | 73.1 (70.2-75.8) |
| **Age, y** |  |  |  |  |  |  |
| 0-5 | 144 (5.7) | 3 (2.1) | 112 (77.8) | 107 (74.3) | 77.2 (70.3-83.3) | 76.7 (69.7-83.0) |
| 6-11 | 242 (9.6) | 23 (9.5) | 231 (95.5) | 217 (89.7) | 91.0 (87.1-94.4) | 90.5 (86.5-94.1) |
| 12-17 | 155 (6.1) | 95 (61.3) | 153 (98.7) | 126 (81.3) | 93.4 (90.3-96.1) | 86.4 (80.2-91.9) |
| 18-24 | 169 (6.7) | 150 (88.8) | 169 (100.0) | 135 (79.9) | 95.0 (93.0-96.9) | 84.0 (77.8-90.1) |
| 25-34 | 267 (10.6) | 239 (89.5) | 263 (98.5) | 209 (78.3) | 94.9 (93.7-95.9) | 78.9 (74.3-83.3) |
| 35-49 | 787 (31.2) | 706 (89.7) | 780 (99.1) | 554 (70.4) | 95.0 (94.2-95.7) | 74.4 (70.5-78.1) |
| 50-64 | 400 (15.9) | 349 (87.2) | 387 (96.8) | 254 (63.5) | 95.4 (94.7-96.1) | 67.7 (62.5-72.6) |
| 65-74 | 183 (7.3) | 173 (94.5) | 179 (97.8) | 92 (50.3) | 95.1 (94.3-95.9) | 55.0 (47.3-62.5) |
| ≥75 | 174 (6.9) | 164 (94.3) | 168 (96.6) | 71 (40.8) | 96.7 (96.2-97.1) | 45.9 (38.3-53.7) |

CrI: credible interval; N: nucleocapsid protein; S: spike protein; SARS-CoV-2: severe acute respiratory syndrome coronavirus 2.
<sup>a</sup> Self-reported having received at least 1 dose of the COVID-19 vaccine before blood sample was drawn.
<sup>b</sup> Serology based on Roche Elecsys anti-SARS-CoV-2 S immunoassay and N immunoassay, respectively.
<sup>c</sup> Seroprevalence estimates reported as % and 95% credible interval, adjusted for test performance of both immunoassays and post-stratified to account for age distribution in the Geneva general population and for household clustering of infection and vaccination. Seroprevalence of any antibodies is based on proportion of participants with any anti-SARS-CoV-2 antibodies, additionally post-stratified to the vaccination data in the general population of Geneva; seroprevalence of infection antibodies is based on proportion of participants who were naturally infected (but could also have been vaccinated).

After accounting for the Geneva population demographics and vaccination status, the overall seroprevalence of anti-SARS-CoV-2 antibodies developed through vaccination and/or infection was 93.8% (95% CrI: 93.1-94.5), with no difference between men and women (Table 1). Estimates varied only slightly across age groups except the youngest; overall seroprevalence among children aged 0-5 years was 77.2% (70.3-83.3) while it was 91.0% (87.1-94.4) among children aged 6-11 years, progressively increasing with age to reach 96.7% (96.2-97.1) among adults aged ≥ 75 years. The overall seroprevalence of infection-induced antibodies was 72.4% (70.0-74.7), also being similar between men and women, but varying considerably across age groups. Among children aged 0-5 years, the estimate was 76.7% (69.7-83.0), while it was highest among children aged 6-11 years at 90.5% (86.5-94.1), markedly decreasing with age and being lowest at 45.9% (38.3-53.7) among adults aged ≥ 75 years (Table 1; Figure 1; Supplementary figure 6; Supplementary table 3). We found no meaningful differences in vaccination rate or seroprevalence estimates according to educational level (Supplementary tables 3-4).

**Figure 1.**
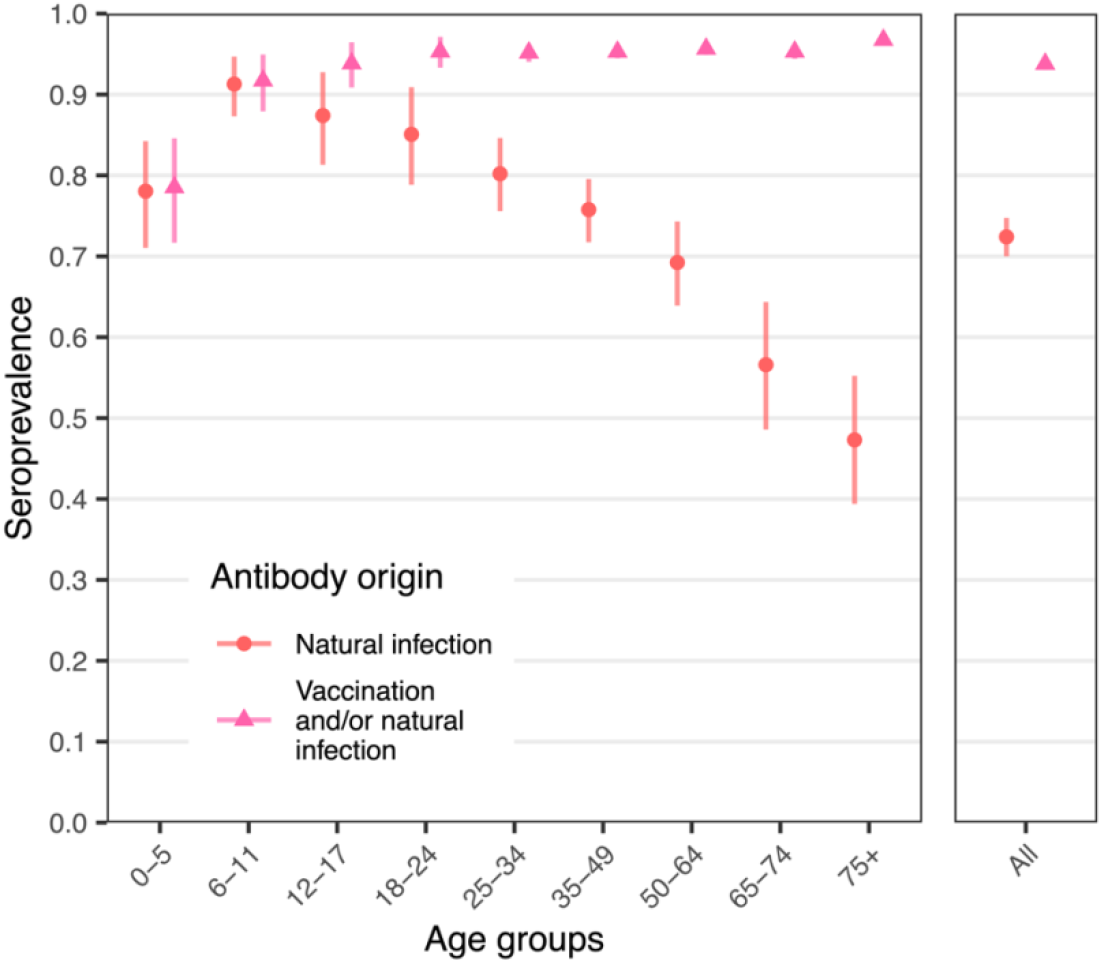
Seroprevalence of anti-SARS-CoV-2 antibodies in the general population of Geneva, Switzerland, April 29^th^ to June 9^th^, 2022 **Seroprevalence estimates in total sample and by age group, serosurvey period and origin of antibody response**. Symbols indicate the antibody origin: *dot* indicates antibodies developed after infection; *triangle* indicates antibodies developed after infection and/or vaccination. Vertical bars represent 95% credible intervals.

### Seroprevalence estimates and determinants of anti-SARS-CoV-2 antibodies neutralizing capacity

Our analytical sample for assessing anti-SARS-CoV-2 neutralization activity included a subset of 1160 participants (Supplementary figure 2), of whom 54.4% were women, 28.2% aged <18 years and 19.3% aged ≥ 65 years (Supplementary figure 4; Supplementary table 5). Overall anti-SARS-CoV-2 seroprevalence estimates on this subsample were similar to those obtained on the main study sample (Supplementary table 6). Distribution of ED50% values obtained on this subsample for all tested SARS-CoV-2 variants using our cell-free surrogate neutralization assay is shown in Supplementary figure 7.

After accounting for the general population demographic and vaccination and infection status distributions, population-level neutralizing capacity was variant-specific (Figure 2; Supplementary figure 8): while 75-80% of the population had neutralizing antibodies against the ancestral D614G, Alpha, and Delta variants, seroprevalence of neutralizing antibodies against all tested Omicron subvariants was lower than 60% (Figure 2; Supplementary table 7; Supplementary figure 8).

**Figure 2.**
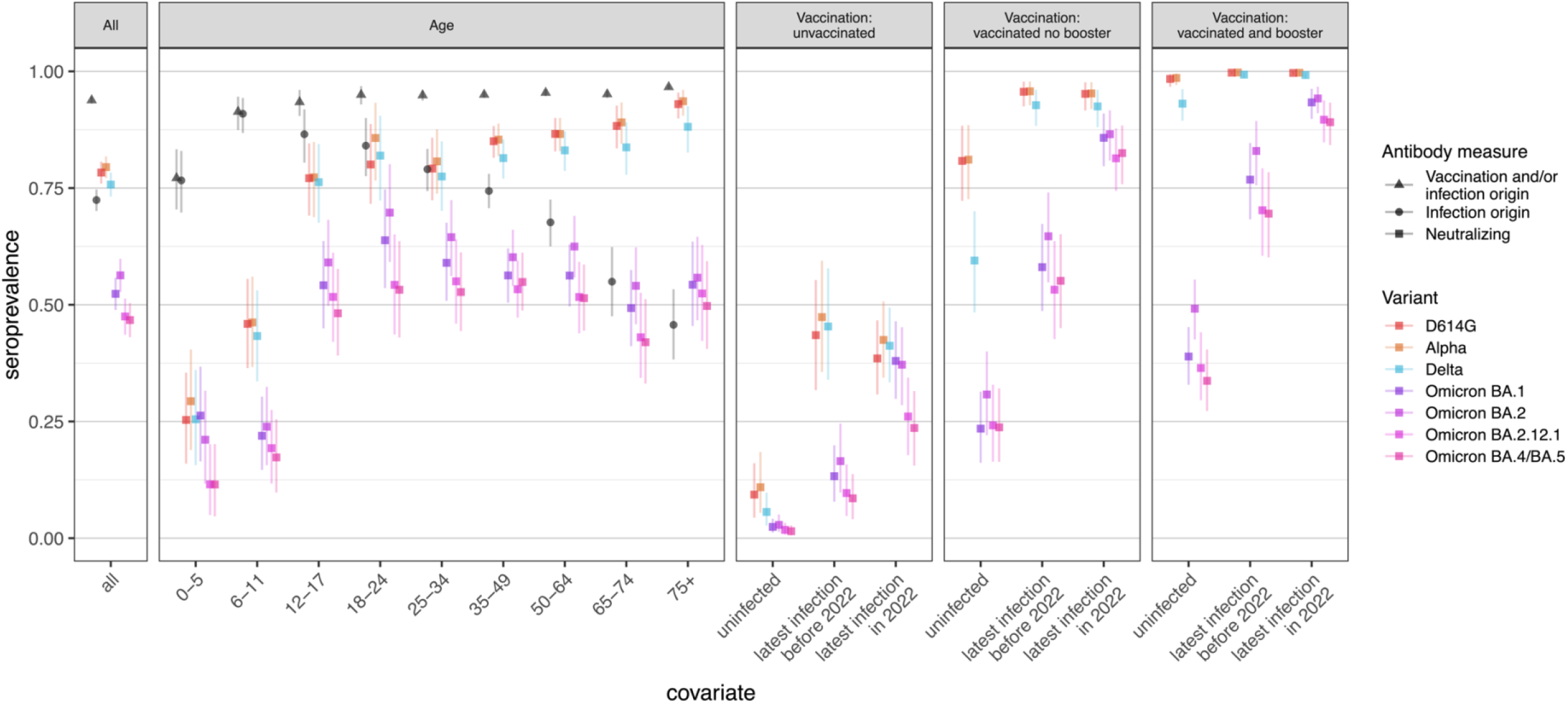
Seroprevalence of neutralizing antibodies against main SARS-CoV-2 variants in the general population of Geneva, Switzerland, April 29^th^ to June 9^th^, 2022 Panels show the effect of covariates tested in the model: age and infection and vaccination statuses (self-reported)—though sex was included as covariate, it showed no apparent effect, so it is excluded here, but estimates are shown in Supplementary figure 8 and Supplementary table 7. Estimates of neutralizing capacity against Beta, Gamma, and Lambda variants are shown in Supplementary figure 8 and Supplementary table 7. Global seroprevalence estimates for anti-S and anti-N antibodies are included in black in the two left panels for comparison purposes. Symbols indicate the antibody origin: *dot* indicates antibodies developed after infection (anti-N); *triangle* indicates antibodies developed after infection and/or vaccination (anti-S); and *square* indicates neutralizing antibodies (cell-free surrogate neutralization assay). Vertical bars represent 95% credible intervals.

Among children aged 0-5 years, estimated seroprevalence of neutralizing antibodies was 29.3% (18.9-40.4) and 26.3% (16.4-36.7) against Alpha and Omicron BA.1, respectively, but 11.5% (4.7-20.2) against Omicron BA.4/BA.5. For children aged 6-11 years, seroprevalence of neutralizing antibodies ranged from 46.2% (36.7-56.0) against Alpha to 17.3% (9.8-25.5) against Omicron BA.4/BA.5. Starting with age 12 years, seroprevalence of neutralizing antibodies against D614G, Alpha and Delta was markedly higher: around 75% among adolescents aged 12-17 years, and around 90% among individuals aged ≥ 75 years. However, seroprevalence of neutralizing antibodies against the Omicron subvariants remained relatively similar, and lower, across these age groups, being 48.2% (39.1-57.7) and 49.7% (40.5-59.3) against BA.4/BA.5 among individuals aged 12-17 years and ≥ 75 years, respectively (Figure 2; Supplementary table 7; Supplementary figure 8).

Seroprevalence of neutralizing antibodies varied considerably according to vaccination and infection statuses (Figure 2; Supplementary table 7; Supplementary figure 8). In general, neutralizing capacity against D614G, Alpha, and Delta variants was substantial (>90%) among vaccinated individuals having received booster vaccination, regardless of infection status. However, among vaccinated individuals without booster vaccination, neutralizing capacity was decreased (reaching 60%) if uninfected. Consistently, in multivariable analyses, having received booster vaccination showed the strongest association with neutralizing capacity; for instance, boosted individuals had 14.2 (95% Cr: 6.4-28.0) times greater odds of having neutralizing antibodies against Delta, and 2.2 (1.4-3.4) times the odds of having antibodies with neutralizing capacity against Omicron BA.4/BA.5, compared with vaccinated individuals without booster vaccination (Table 2). Regarding infection, compared with individuals last infected before 2022, those infected in 2022 had more than three times greater odds of having neutralizing antibodies against the Omicron subvariants. Finally, regardless of variant, unvaccinated individuals had substantially lower odds of having neutralizing antibodies (Table 2).

**Table 2.**
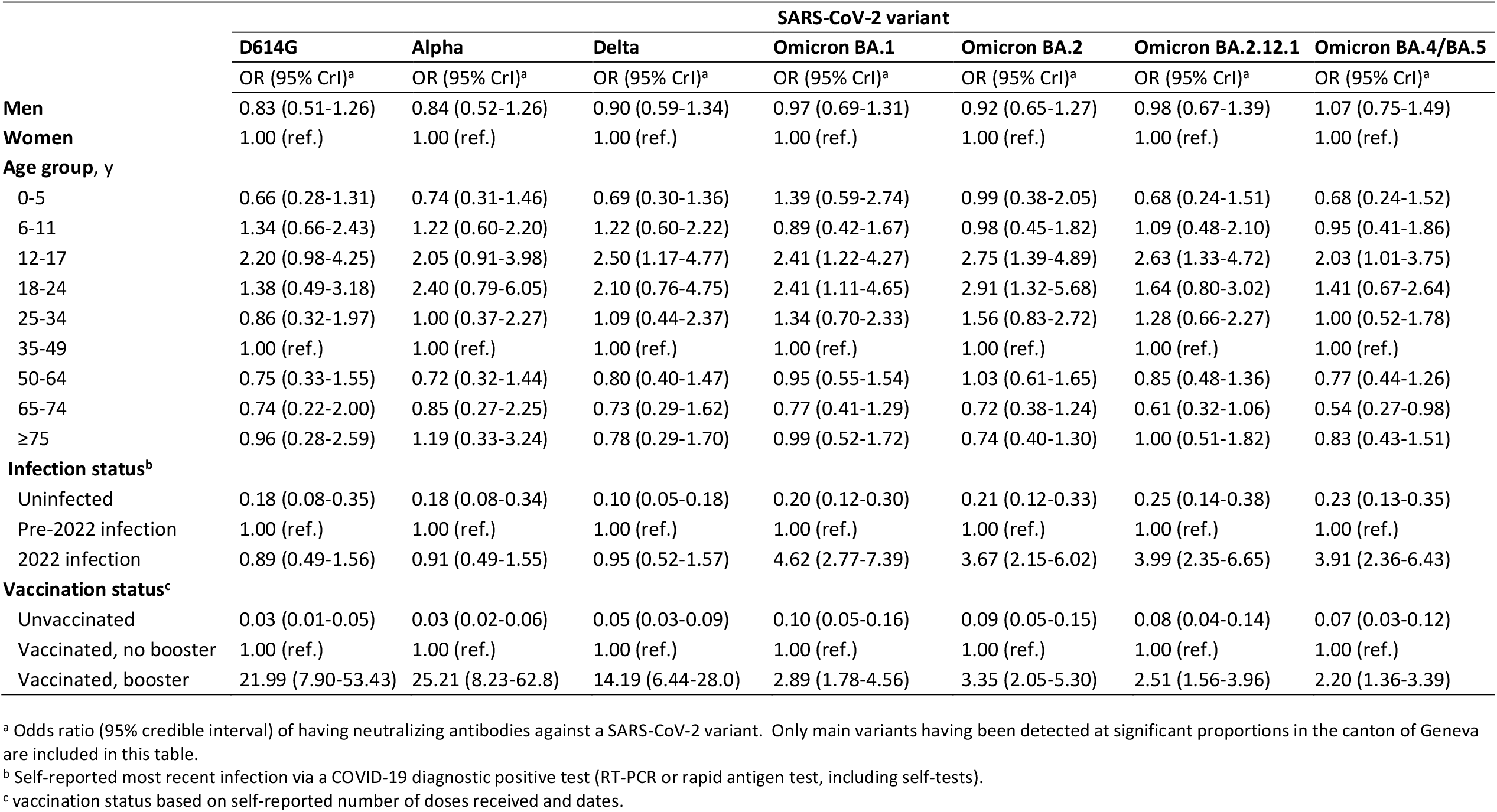
Association between individuals’ attributes and neutralizing capacity against main SARS-CoV-2 variants in the general population of Geneva, Switzerland, April 29^th^ to June 9^th^, 2022

|  | SARS-CoV-2 variant |  |  |  |  |  |  |
| --- | --- | --- | --- | --- | --- | --- | --- |
|  | D614G | Alpha | Delta | Omicron BA.1 | Omicron BA.2 | Omicron BA.2.12.1 | Omicron BA.4/BA.5 |
|  | OR (95% CrI)<sup>a</sup> | OR (95% CrI)<sup>a</sup> | OR (95% CrI)<sup>a</sup> | OR (95% CrI)<sup>a</sup> | OR (95% CrI)<sup>a</sup> | OR (95% CrI)<sup>a</sup> | OR (95% CrI)<sup>a</sup> |
| **Men** | 0.83 (0.51-1.26) | 0.84 (0.52-1.26) | 0.90 (0.59-1.34) | 0.97 (0.69-1.31) | 0.92 (0.65-1.27) | 0.98 (0.67-1.39) | 1.07 (0.75-1.49) |
| **Women** | 1.00 (ref.) | 1.00 (ref.) | 1.00 (ref.) | 1.00 (ref.) | 1.00 (ref.) | 1.00 (ref.) | 1.00 (ref.) |
| **Age group, y** |  |  |  |  |  |  |  |
| 0-5 | 0.66 (0.28-1.31) | 0.74 (0.31-1.46) | 0.69 (0.30-1.36) | 1.39 (0.59-2.74) | 0.99 (0.38-2.05) | 0.68 (0.24-1.51) | 0.68 (0.24-1.52) |
| 6-11 | 1.34 (0.66-2.43) | 1.22 (0.60-2.20) | 1.22 (0.60-2.22) | 0.89 (0.42-1.67) | 0.98 (0.45-1.82) | 1.09 (0.48-2.10) | 0.95 (0.41-1.86) |
| 12-17 | 2.20 (0.98-4.25) | 2.05 (0.91-3.98) | 2.50 (1.17-4.77) | 2.41 (1.22-4.27) | 2.75 (1.39-4.89) | 2.63 (1.33-4.72) | 2.03 (1.01-3.75) |
| 18-24 | 1.38 (0.49-3.18) | 2.40 (0.79-6.05) | 2.10 (0.76-4.75) | 2.41 (1.11-4.65) | 2.91 (1.32-5.68) | 1.64 (0.80-3.02) | 1.41 (0.67-2.64) |
| 25-34 | 0.86 (0.32-1.97) | 1.00 (0.37-2.27) | 1.09 (0.44-2.37) | 1.34 (0.70-2.33) | 1.56 (0.83-2.72) | 1.28 (0.66-2.27) | 1.00 (0.52-1.78) |
| 35-49 | 1.00 (ref.) | 1.00 (ref.) | 1.00 (ref.) | 1.00 (ref.) | 1.00 (ref.) | 1.00 (ref.) | 1.00 (ref.) |
| 50-64 | 0.75 (0.33-1.55) | 0.72 (0.32-1.44) | 0.80 (0.40-1.47) | 0.95 (0.55-1.54) | 1.03 (0.61-1.65) | 0.85 (0.48-1.36) | 0.77 (0.44-1.26) |
| 65-74 | 0.74 (0.22-2.00) | 0.85 (0.27-2.25) | 0.73 (0.29-1.62) | 0.77 (0.41-1.29) | 0.72 (0.38-1.24) | 0.61 (0.32-1.06) | 0.54 (0.27-0.98) |
| ≥75 | 0.96 (0.28-2.59) | 1.19 (0.33-3.24) | 0.78 (0.29-1.70) | 0.99 (0.52-1.72) | 0.74 (0.40-1.30) | 1.00 (0.51-1.82) | 0.83 (0.43-1.51) |
| **Infection status<sup>b</sup>** |  |  |  |  |  |  |  |
| Uninfected | 0.18 (0.08-0.35) | 0.18 (0.08-0.34) | 0.10 (0.05-0.18) | 0.20 (0.12-0.30) | 0.21 (0.12-0.33) | 0.25 (0.14-0.38) | 0.23 (0.13-0.35) |
| Pre-2022 infection | 1.00 (ref.) | 1.00 (ref.) | 1.00 (ref.) | 1.00 (ref.) | 1.00 (ref.) | 1.00 (ref.) | 1.00 (ref.) |
| 2022 infection | 0.89 (0.49-1.56) | 0.91 (0.49-1.55) | 0.95 (0.52-1.57) | 4.62 (2.77-7.39) | 3.67 (2.15-6.02) | 3.99 (2.35-6.65) | 3.91 (2.36-6.43) |
| **Vaccination status<sup>c</sup>** |  |  |  |  |  |  |  |
| Unvaccinated | 0.03 (0.01-0.05) | 0.03 (0.02-0.06) | 0.05 (0.03-0.09) | 0.10 (0.05-0.16) | 0.09 (0.05-0.15) | 0.08 (0.04-0.14) | 0.07 (0.03-0.12) |
| Vaccinated, no booster | 1.00 (ref.) | 1.00 (ref.) | 1.00 (ref.) | 1.00 (ref.) | 1.00 (ref.) | 1.00 (ref.) | 1.00 (ref.) |
| Vaccinated, booster | 21.99 (7.90-53.43) | 25.21 (8.23-62.8) | 14.19 (6.44-28.0) | 2.89 (1.78-4.56) | 3.35 (2.05-5.30) | 2.51 (1.56-3.96) | 2.20 (1.36-3.39) |

<sup>a</sup> Odds ratio (95% credible interval) of having neutralizing antibodies against a SARS-CoV-2 variant. Only main variants having been detected at significant proportions in the canton of Geneva are included in this table.
<sup>b</sup> Self-reported most recent infection via a COVID-19 diagnostic positive test (RT-PCR or rapid antigen test, including self-tests).
<sup>c</sup> vaccination status based on self-reported number of doses received and dates.

## DISCUSSION

This serosurvey found that by April-June 2022, 93.8% of the Geneva population had developed antibodies against SARS-CoV-2 after vaccination and/or infection. Yet the proportion of the population with neutralizing antibodies varied considerably across SARS-CoV-2 variants, from more than three quarters against Alpha to less than half against the currently circulating Omicron BA.5 subvariant, with particularly lower proportions in children <12 years and unvaccinated individuals who were last infected before Omicron became dominant.

The seroprevalence of total antibodies estimated in this study is slightly lower than the 97-98% reported in the United Kingdom by the beginning of May 2022 (28). We also found that 72.4% of the population had been infected—a 42.5 percentage point increase from the 29.9% seroprevalence reported by June-July, 2021 (20) (Supplementary table 8). This important increase in seroprevalence of infection-induced antibodies within a 11-month period was largest among children aged 0-5 years (55.8 percentage point increase) and 6-11 years (59.5 percentage point increase), indicating that the Delta-dominant and notably the Omicron-dominant pandemic waves in Geneva particularly affected children (29), as observed in other countries (28,30). Conversely, this increase in infection-induced antibodies does not appear to have translated to a corresponding level of neutralizing capacity; for instance, while three quarters of children aged 0-5 years had developed antibodies through infection, only around one in four had neutralizing antibodies against the Delta or Omicron BA.1 or BA.2 variants. This proportion was only slightly higher among the 6-11 years old even though nine in ten children in this age group had infection-induced antibodies. These findings indicate that most children, not having received the COVID-19 vaccine and first becoming infected by Delta or Omicron BA.1 or BA.2, did not develop neutralizing capacity against earlier variants and only limited neutralizing capacity against currently circulating BA.5 subvariant.

Our model suggests that this discrepancy can be explained by the significantly lower vaccination rates in these age groups alone, without requiring age-specific effects (95% CrI of odds ratios for all variants and for all age groups cover 1, except for the 12-17 years age group, which had higher odds of neutralization). We however note that most infected-only participants in our sample were children under the age of 12 years, which therefore limits the power to identify age-specific effects within the analysis. Although antibodies in children seem to have a lower neutralization efficacy, their immune response has been observed to resemble that of adults in cases of mild COVID-19, but to differ in moderate to severe COVID-19 and multisystem inflammatory syndrome (31), including a robust mucosal response to the SARS-CoV-2 virus with high levels of interferon and a different T-cell response (31,32). While data on these other immunity components were not available in our study, children-specific immune response may explain the consistently lower levels of severe COVID-19, hospitalizations, and death observed among children, despite much higher levels of infections during the Delta- and Omicron-driven waves (20).

We found little age differences in seroprevalence of total antibodies among adults, in contrast to what we observed in our previous serosurvey (20), likely reflecting the fact that for adults vaccination has since been widely available. Simultaneously, seroprevalence of infection-induced antibodies differed markedly among age groups, peaking for the 6-11 years age group and reducing gradually with increasing age, reflecting the pattern of age-related infection risk observed across pandemic waves, preventive measures and behavior, as well as the earlier vaccination availability and higher uptake in older people (1,3,20,30).

The finding that neutralizing capacity against the D614G, Alpha, and Delta variants was reduced among individuals infected in 2022 compared with those last infected before 2022 is in line with reports of reduced overall neutralizing capacity against pre-Omicron variants after infection by Omicron (8,13,16). Notably, we also found that having received booster vaccination was associated with increased neutralizing capacity against all variants, including Omicron subvariants, in agreement with previous reports (8,9,13,16,17). Finally, we found that being vaccinated (with or without booster) and infected in 2022, a period when Omicron was almost exclusively circulating in Geneva, was associated with increased neutralizing capacity against all Omicron subvariants, in line with previous reports (9,13,16,17,33). In general, a combination of vaccination (with booster) and recent infection appeared to confer the highest level of neutralizing antibodies against each variant, including Omicron subvariants.

### Implications for public health and clinical practice

Our findings show that, while most of the population have developed anti-SARS-CoV-2 antibodies through infection and/or vaccination, less than half have neutralizing antibodies against the highly contagious, currently circulating Omicron BA.5 variant, including only one in four children aged <12 years. A similar pattern of total antibodies and variant-specific neutralizing antibodies in the population is likely to be present in other settings that have experienced the same successive variant-driven pandemic waves as Geneva, Switzerland. The highest level of neutralizing capacity against each variant was observed among vaccinated individuals who had received a booster dose, indicating that vaccine-induced antibodies confer substantial neutralizing capacity against pre-Omicron variants, while also maximizing neutralizing capacity against Omicron subvariants. At the same time, not surprisingly, we observed a reduced neutralizing capacity against Omicron subvariants relative to pre-Omicron variants. This suggests that updated vaccines specifically targeting the Omicron lineage may be beneficial in containing the spread of infections and their consequent health and socio-economic burden (1).

Our findings also revealed that, while less than half of the population show neutralizing capacity against the currently dominant Omicron subvariants, a substantial proportion have neutralizing capacity against less common variants, including Beta, Gamma, and Lambda (Supplementary table 7, Supplementary figure 7). Since future VOCs may develop from or share structural characteristics with less frequent variants, monitoring the level of neutralizing capacity against them in the population may help in building scenarios for future pandemic waves.

### Strengths and limitations

This study benefits from several strengths, including the large representative sample, the recent recruitment time-frame post-Omicron BA.1 and BA.2-driven pandemic waves, the measurement of antibodies against both the SARS-CoV-2 S and N proteins as well as neutralizing antibodies against 10 SARS-CoV-2 variants/subvariants, and a robust modelling framework. We also acknowledge several limitations. First, like most serosurveys (34), the sample only included formal residents of the canton and had a higher proportion of individuals with tertiary education than in the general population (Supplementary table 1), although we did not observe differences in seroprevalence estimates across education levels. Second, data on infection dates was self-reported by the participants at the moment of the blood drawing and only the latest infection was included in the analysis. Third, the cut-off value for neutralizing activity was defined using pre-pandemic sera from adult donors only— no children samples were included in the validation study, as previously reported (23). While the level of neutralizing antibodies has been shown to be a strong marker of immune protection, it does not fully describe it, especially among children whose immune response differs from that of adults (31,32). Lastly, due to lack of data, we did not include in our modelling framework indicators that have been shown to influence neutralizing capacity, including severity and duration of symptoms, number of infections, and interval between last infection/vaccination and blood sampling (9–11,35,36).

## CONCLUSION

This study provides up-to-date seroprevalence estimates of anti-SARS-CoV-2 antibodies in a representative sample of the general population 5 months after Omicron became the dominant circulating SARS-CoV-2 variant in Geneva, Switzerland. It shows that while most of the population (notably ≥ 12 years of age) have neutralizing antibodies against pre-Omicron variants, the seroprevalence of antibodies with neutralizing capacity against the currently circulating and highly contagious Omicron BA.5 subvariant is low. Our findings suggest that the mass vaccination of older individuals, as well as other preventive measures and behaviors, may have protected them from infection during the Delta- and Omicron BA.1- and BA.2-driven waves. They also show that the highest level of neutralizing capacity against most VOCs is attained through hybrid immunity combining vaccination, notably including a booster dose, and recent infection. As new variants emerge driving new pandemic waves, having up-to-date snapshots of the immune landscape of the population can help develop rational risk mitigation strategies.

## Data Availability

Data can be made available upon submission of a data request application to the corresponding author. Biological material can be reused for further studies upon approval by the cantonal ethics commission of the state of Geneva.

## Acknowledgments

We warmly thank the HEdS students Léa Baettig, Jessica Chavet, Noé Kummer, Manuel Lobato Sineiro and Reza Nazari and the nurses Nassima Sadadou Djouder, Manon Ladouce, Marie Le Belz, Celine Breuil and Marion Figini for their passionate work during this study. We thank Florence Pojer, Kelvin Lau and the Protein Production and Structure Core Facility team at EPFL for the tremendously efficient Spike variants production, and Vanessa Genet at EPFL for her precious technical help. We are grateful to the Hôpital des enfants des HUG, the Hôpital de La Tour, the Clinique and Permanence d’Onex and the Centre Médical du Lignon for allowing us to use their premises for the recruitment of participants, and to Dr Cyril Sahyoun and his team for our enriching collaboration aimed at easing the experience of children during blood sampling. Finally, we are deeply grateful to all the participants for their interest and invaluable contribution to the study.

