## Supplementary file for "Seroprevalence of anti-SARS-CoV-2 antibodies and cross-variant neutralization capacity after the Omicron BA.2 wave in Geneva, Switzerland"

**Supplementary information S1.** Questions given to participants and used in the present study

| Question | Answer choices |
|---|---|
| What is your date of birth? | day / month / year |
| What is your sex? | Male / Female / Other |
| ***Vaccination*** | |
| Have you received the first dose of the vaccine against COVID-19? | Yes / No |
| Have you received the second dose of the vaccine against COVID-19? | Yes / No<br>If yes, date (month / year) |
| Have you received the "booster" dose of the vaccine against COVID-19? | Yes / No<br>If yes, date (day / month / year) |
| Since the beginning of the pandemic, have you had a confirmed COVID-19 diagnosis, meaning having a positive COVID-19 test from a nasal/throat swab, either RT-PCR or rapid antigenic test, including self-test? | Yes / No<br>If yes, date (month / year) [for each positive test] |
| ***Educational level*** | |
| What is the highest level diploma/certification that you have obtained? | - None<br>- Primary school and/or orientation cycle<br>- Secondary education – *Maturité*/High School<br>- Professional training – Certified apprenticeship (*CFC*)<br>- Professional training – Non-certified apprenticeship<br>- Professional training – Higher professional degrees (post-*CFC*)<br>- Tertiary education – Bachelor/Master degree<br>- Tertiary education – Doctorate/PhD degree<br>- Other |

**Supplementary information S2. Overview of Statistical Frameworks**

### S2.1 Antibody presence and origin

Our first aim was to infer the proportion of the population having any antibodies against SARS-CoV-2, as well as the proportion of those who acquired antibodies through natural infection as opposed to vaccination-only. We did so following our previous work by modelling jointly the antibody response measured by the Roche-N and Roche-S immunoassays together with participants' responses to a vaccination questionnaire (1). Population-level estimates of antibody seroprevalence of any or infection origin were then obtained using demographic statistics on the population of Geneva, as well as official statistics on age-specific vaccination rates in the population (Supplementary table 2). Post-stratification therefore allows to correct for the demographic biases of our sample.

We disentangled natural infection from vaccination antibody responses using the fact that the only available vaccines in Switzerland to date—the mRNA-1273 from Moderna/US NIAID (2), the mRNA-BNT162b2/Comirnaty from Pfizer/BioNTech (3), and the Janssen Ad26.COV2.S COVID-19 vaccine (4)—elicit a response exclusively to the S protein of SARS-CoV-2, as opposed to natural infections which typically elicit a response to both the N and S virus proteins. We expanded previous Bayesian modelling frameworks used for seroprevalence estimates that account for demographic parameters (sex and age), test performance and household infection clustering (5,6). The main difference with our previous study is that here we considered that the probability of vaccination is independent from the probability of infection, which was not the case by June-July 2021 due to Geneva's vaccination policy (1).

#### *S2.1.1 Multinomial response model*

We modeled the Roche-S and Roche-N tests results for participant $i$, $x_i$, consisting of one of four possible outcome combinations $[n_{S^+N^+}, n_{S^-N^+}, n_{S^+N^-}, n_{S^-N^-}]$ (+ indicates antibody presence; $-$ indicates absence) with $x_i \in \{[1,0,0,0],[0,1,0,0],[0,0,1,0],[0,0,0,1]\}$ using a multinomial distribution parametrized by parameter vector $\pi_i = [\pi_i^{++}, \pi_i^{-+}, \pi_i^{+-}, \pi_i^{--}]$, where $\pi^{lm}$ is the probability of having Roche-S test result $l$ and Roche-N result $m$, accounting for both the underlying probability of each antibody status $p^{\pm/\pm}$, and test sensitivity, $\theta^+$and specificity, $\theta^-$:

$$x_i \sim \text{Multinomial}(\pi_i)$$

$$\pi_i^{++} = \theta_S^+\theta_N^+p_i^{++} + (1-\theta_S^-)\theta_N^+p_i^{-+} + \theta_S^+(1-\theta_N^-)p_i^{+-} + (1-\theta_S^-)(1-\theta_N^-)p_i^{--},$$

$$\pi_i^{-+} = (1-\theta_S^+)\theta_N^+p_i^{++} + \theta_S^-\theta_N^+p_i^{-+} + (1-\theta_S^+)(1-\theta_N^-)p_i^{+-} + \theta_S^-(1-\theta_N^-)p_i^{--},$$

$$\pi_i^{+-} = \theta_S^+(1-\theta_N^+)p_i^{++} + (1-\theta_S^-)(1-\theta_N^+)p_i^{-+} + \theta_S^+\theta_N^-p_i^{+-} + \theta_S^-(1-\theta_N^-)p_i^{--},$$

$$\pi_i^{--} = (1-\theta_S^+)(1-\theta_N^+)p_i^{++} + \theta_S^-(1-\theta_N^+)p_i^{-+} + (1-\theta_S^+)\theta_N^-p_i^{+-} + \theta_S^-\theta_N^-p_i^{--}.$$

The underlying probability of antibody status accounts both for the probability of natural infection $\lambda_i$ and vaccination status, $v_i \in \{0,1\}$. Following previous modeling frameworks(5,6), we modeled the probability of natural infection as a function of sex and age category, accounting for household infection clustering through a random effect, $\alpha_h$:

$$logit(\lambda_i) = \alpha_h + \mathbf{X_i}\boldsymbol{\beta}$$

$$\alpha_h \sim Normal(0, \sigma_h^2)\,,$$

where $\mathbf{X}_i$ is the matrix of covariates, and $\boldsymbol{\beta}$ the vector of regression coefficients. The probabilities of antibody status are then given by:

$$p_i^{++} = \gamma^{++}\lambda_i + \gamma^{-+}v_i$$

$$p_i^{-+} = \gamma^{-+}\lambda_i\ (1-v_i)$$

$$p_i^{+-} = (1-\lambda_i)v_i + \gamma^{+-}\lambda_i$$

$p_i^{--} = 1 - v_i(1 - \lambda_i) - \lambda_i$,

where $\gamma^{++}, \gamma^{-+}, \gamma^{+-}$ are the conditional probability of having $S^+N^+, S^-N^+, S^+N^-$ responses, respectively, upon natural infection, $v_i$ is the probability of having a vaccine-induced $S^+$ response as a function of the conditional probability of antibody response upon infection $\eta_i$, $v_i = \eta_i \times v_i$.

#### *S2.1.2 Vaccination*

To obtain population-level seroprevalence estimates, we also modeled the proportion of vaccinated individuals in each sex/age class following the approach used for natural infection:

$v_i \sim \text{Bernoulli}(\phi_i)$ ,

$logit(\phi_i) = \alpha_{v,h} + \mathbf{X}_i\boldsymbol{\beta_v}$,

$\alpha_{v,h} \sim \text{Normal}(0, \sigma_v^2)$.

#### *S2.1.3 Diagnostic test performance*

The individual performance of both N and S tests was incorporated hierarchically following Gelman & Carpenter (7). The sensitivity, $\theta^+$, was determined using $n^+$ RT-PCR positive controls from a laboratory validation study (8), of which $x^+$ tested positive. The specificity, $\theta^-$, was determined using $n^-$ pre-pandemic negative controls, of which $x^-$ tested positive. For the Roche N test, these values were modulated by data in Ainsworth et al. (9). For the Roche S test, the laboratory study data were modulated by those available on the Roche website (last accessed July 20, 2022: https://diagnostics.roche.com/global/en/products/params/elecsys-anti-sars-cov-2-s.html).

#### *S2.1.4 Priors*

We followed a similar setting of the priors on the tests' sensitivity and specificity as Gelman & Carpenter (7). For study j, the specificity $\theta_j^-$ and sensitivity $\theta_j^+$ were drawn from normal distributions on the log odds scale:

$logit(\theta_j^-) \sim \text{Normal}(\mu_{\theta-}, \sigma_{\theta-})$

$logit(\theta_j^+) \sim \text{Normal}(\mu_{\theta+}, \sigma_{\theta+})$.

Hyperparameters $\mu_z$ and $\sigma_z$ for $z \in (\theta^-, \theta^+)$ follow, on the logit scale, normal distributions $\mu_z \sim \mathcal{N}(4,2)$ and positive half-normals $\sigma_z \sim \mathcal{N}^+(0,1)$, respectively. These priors on test performance were identical for both the Roche S and Roche N tests.

We used standard normal $\mathcal{N}(0,1)$ priors for the logistic regression coefficients for infection $\boldsymbol{\beta}$. For coefficients of vaccination $\boldsymbol{\beta_v}$ we used $\mathcal{N}(0,5)$. The priors for the means of the household random effects $\alpha_h$ and $\alpha_{h,v}$, followed standard normal, and for standard deviations of the household random effects were positive half-normals, $\sigma_h \sim \mathcal{N}^+(0,2)$ and $\sigma_v \sim \mathcal{N}^+(0,2)$. We used a Dirichlet prior on the conditional probability of having $S^+N^+, S^-N^+, S^+N^-$ responses upon natural infection, $\gamma^{++}, \gamma^{-+}, \gamma^{+-}, \gamma \sim \text{Dir}(10,1,1)$, to highly favor production of both anti-S and anti-N antibodies upon infection. Finally, we put a strong prior on the conditional probability of antibody response after vaccination $\eta_i \sim \text{Beta}(10, 0.1)$.

#### *S2.1.5 Implementation*

The model was coded in the probabilistic programming language Stan (10) using the Rstan package (11). R version 4.1 (12) was used for data analysis. Four chains were run with 1500 iterations each, 250 of which were warmup, to give a total of 5000 posterior samples. Convergence was assessed by checking that $\widehat{R} \approx 1$, that the effective sample size was reasonable for all parameters, and visually using shinystan (13) diagnostics checks.

#### S2.2 Neutralization capacity modeling

The two main aims of the analysis were first to assess the impact of age, sex, and vaccination and infection status on the variant-specific neutralization capacity, and second to produce population-level estimates through post-stratification.

##### *S2.2.1 Model description*

We developed a logistic regression model accounting for participant characteristics as well as neutralization test performance. The serum sample of participant $i$ was considered to have neutralization capacity if it's ED50 value was above 50 (14). We modeled the presence of neutralization ($p_i \in \{0,1\}$) as a function of the participant's characteristics as:

$$y_i \sim Bernoulli\big(\theta^{+} p_i + (1-\theta^{-})(1-p_i)\big)$$

$$\text{logit}(p_i) = \alpha + \beta_1 \text{ sex}_i + \beta_2 \text{ age}_i + \beta_3 \text{ latest\_inf}_i + \beta_4 \text{ vacc\_status}_i$$

$$n_{tp} \sim Binomial(N^{+}, \theta^{+})$$

$$n_{fp} \sim Binomial(N^{-}, \theta^{-})$$

where $\theta^{+}$ and $\theta^{-}$ are the pseudo-neutralization test's sensitivity and specificity which are informed by the number of true ($n_{tp} = 118$) and false positives ($n_{fp} = 0$) among the validation set of positive ($N^{+} = 122$) and negative ($N^{-} = 84$) controls in Fenwick et al. (14). Latest infection status indicates whether participants were infected by a pre-Omicron SARS-CoV-2 variant (infection prior to January 2022 as Omicron became dominant in late December 2021 in Geneva, Supplementary Figure 6), an Omicron variant (infection in 2022), or uninfected at the time of blood draw.

Infection status was assessed based on anti-N and anti-S Roche serology, and latest infection date was based on self-reported dates of positive tests (RT-PCR or rapid antigenic test, including self-test). Among participants with known infection (positive anti-N or positive anti-S in the absence of vaccination), 41% (341/837) did not report any positive tests. The unknown infection date (before January 2022 or during 2022) for these participants was marginalized out within the modeling framework using prior probabilities of infection before or after January 2022 (non-Omicron vs. Omicron infections). Prior probabilities were determined based on the difference between the age-specific cumulative infection probabilities in this work with those of our previous estimates in June-July 2021 (Supplementary Figure 6) (1). These differences in cumulative attack rates account both for the Delta-driven and Omicron-driven waves in Geneva (Supplementary Figure 6). To compute the prior probabilities of non-Omicron vs. Omicron infections we therefore assumed that approximately 80% of the cumulative attack rate between the two serosurveys was due to Omicron variants based on the ratio of reported cases in the Delta vs. Omicron periods (31953 cases were reported between July 15th and December 25th, 2021 when the Delta variant were dominant, and 123554 cases between December 26th, 2021 and when the serosurvey started on April 29th, 2022 when Omicron subvariants were dominant).

Parameter priors were set following the approach for modeling antibody presence described in section S2.1.4 using $Normal(0,1)$ priors on the intercept and regression parameters.

##### *S2.2.2 Alternative models*

To account for possible effect modification of vaccination effect by infection status, we also fit a model with interaction terms between vaccination status and latest infection status. Over-dispersion of neutralization capacity conditional on participant characteristics was tested through a participant-level additive random effect on the logit-scale. Finally we also tested whether there was support for differential pseudo-neutralization test performance for children (< 12 years old) and other participants given that the test in Fenwick et al. (14) was validated on adult samples only. This was done following a hierarchical model on test performance as done for accounting for multiple validation sets to infer antibody presence in this work and elsewhere (1).

#### *S2.2.3 Model selection*

Model selection was performed through Bayesian estimates of leave-one-out cross-validation error (15), as implemented in the loo R package (16). We accounted for model comparison uncertainty by retaining the simplest model among the set of models with an estimated pairwise log-predictive density difference larger than 2 standard errors. Across all variants, this model selection procedure retained the additive logistic model without over-dispersion and same test performance for children and other participants.

#### *S2.2.4 Post-stratification*

Following the methods described for antibody presence, we obtained population-level estimates of neutralization capacity through post-stratification based on variant-specific parameters estimates from the additive logistic models. For vaccination we used the official cantonal statistics on vaccination as done for antibody presence (Supplementary Table 2). For latest infection status we used our estimates of cumulative attack rates based on anti-N positive serology as described in section S.2.1 (Supplementary Figure 6). Stratification across combinations of latest infection status and vaccination status were based on observed age-specific frequencies in our full analytical sample, scaled to match cantonal vaccination data and our inferred total infection prevalence.

**Supplementary figure 1.** Timeline of serosurveys and vaccination rollout in Geneva, Switzerland

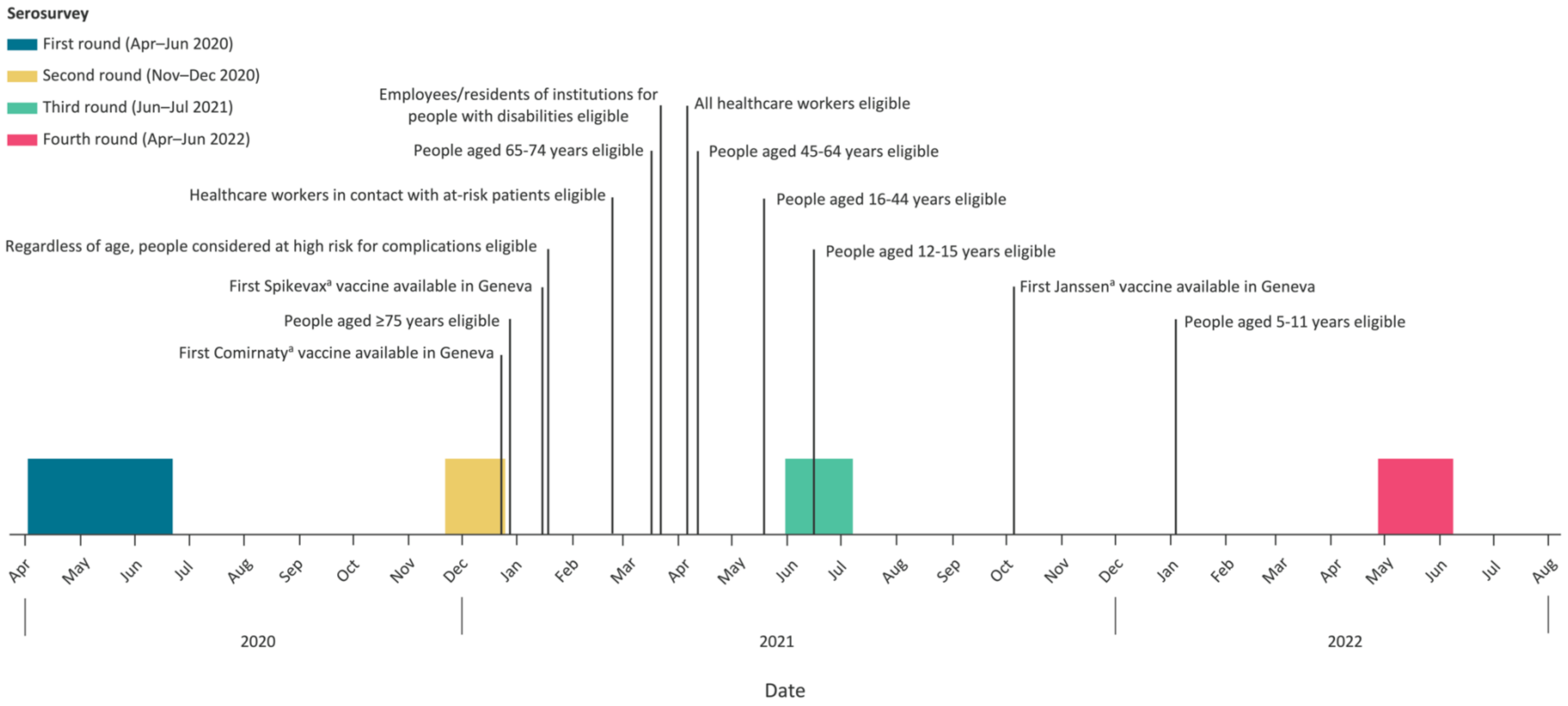


[a] During the course of the first three serosurveys (April 2020–June 2022), the two vaccines approved for use in Switzerland were Comirnaty (BNT162b2, mRNA BioNTech-Pfizer, Mainz, Germany/New York City, United States) and Spikevax (mRNA-1273, Moderna, Cambridge, US); by the fourth serosurvey, the Janssen COVID-19 vaccine (Ad26.COV-S recombinant, Janssen-Cilag International, Bersee, Belgium) was also approved. Eligibility for vaccination allowed individuals to register for an appointment to receive the first vaccine dose.

**Supplementary figure 2.** Participants recruitment and inclusion into analytical samples

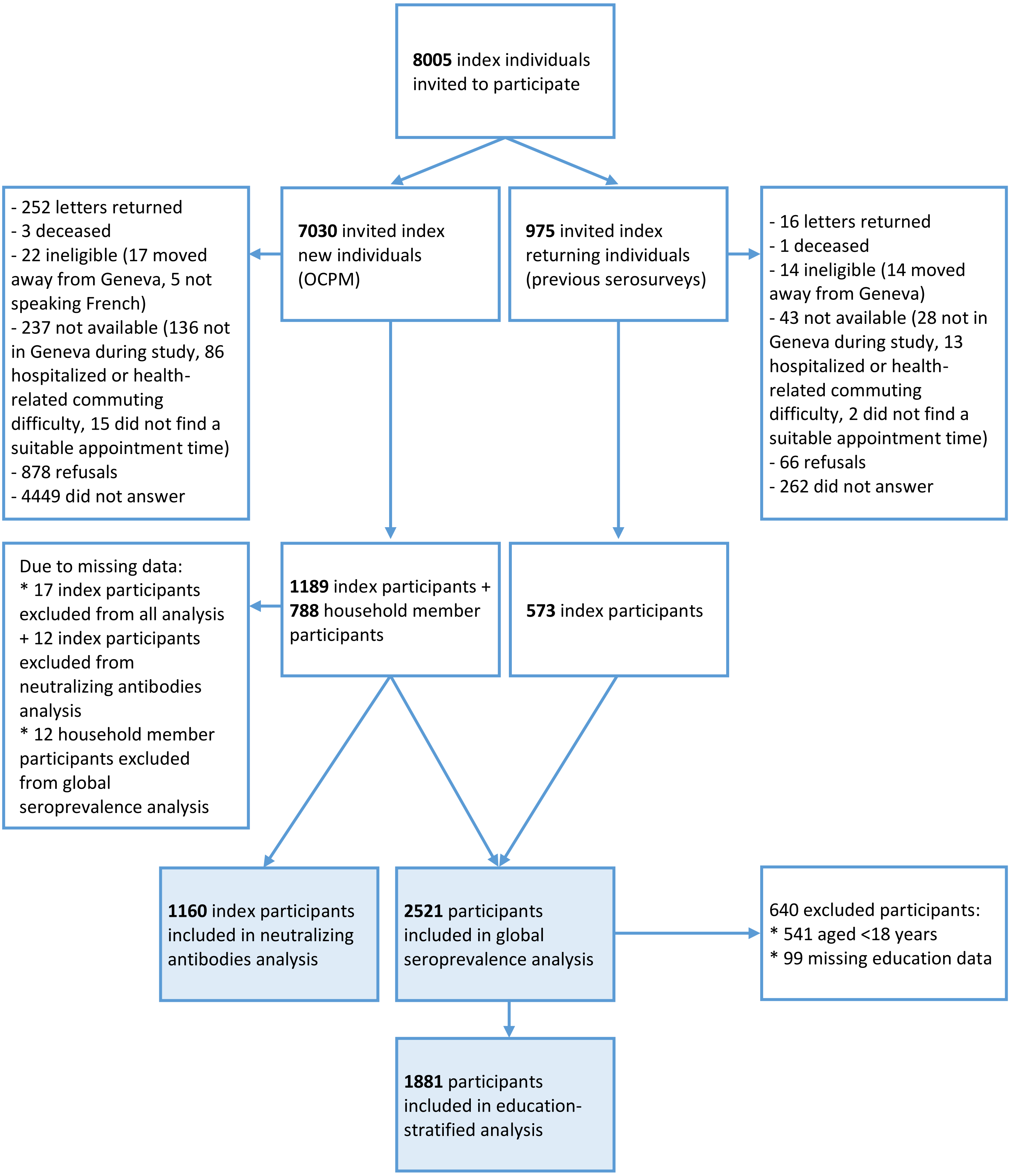

**Supplementary figure 3.** Reasons for refusal to participate to the study as provided by invited individuals

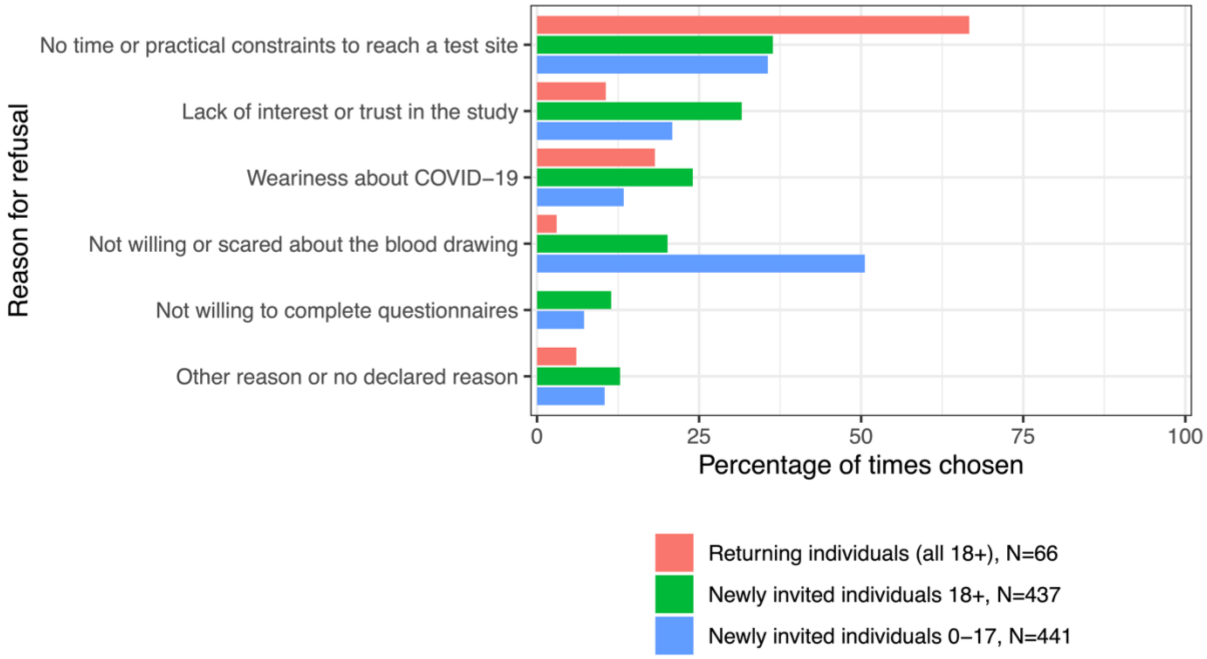


Excluding those ineligible and not available, 17% of the remaining invited individuals (944/5655) provided us with one or more reasons for their refusal to participate to the study (either by direct contact by phone or email or by completing the pseudo-anonymized reply form provided with the reminder letter/email). On the same form, invited individuals were asked, if willing, to indicate their COVID-19 vaccination status and education level (as highest level diploma obtained). The study sample had a higher proportion of individuals with tertiary education as well as a higher proportion of vaccinated individuals than this "refusing" sample; in line with what we observed when comparing the study sample with the official numbers from the canton of Geneva.
Ages "18+" or "0-17" refer to age of invited index individual, in years.

**Supplementary figure 4.** Comparison of age and sex composition of study samples (bars) and the Geneva population (dots)

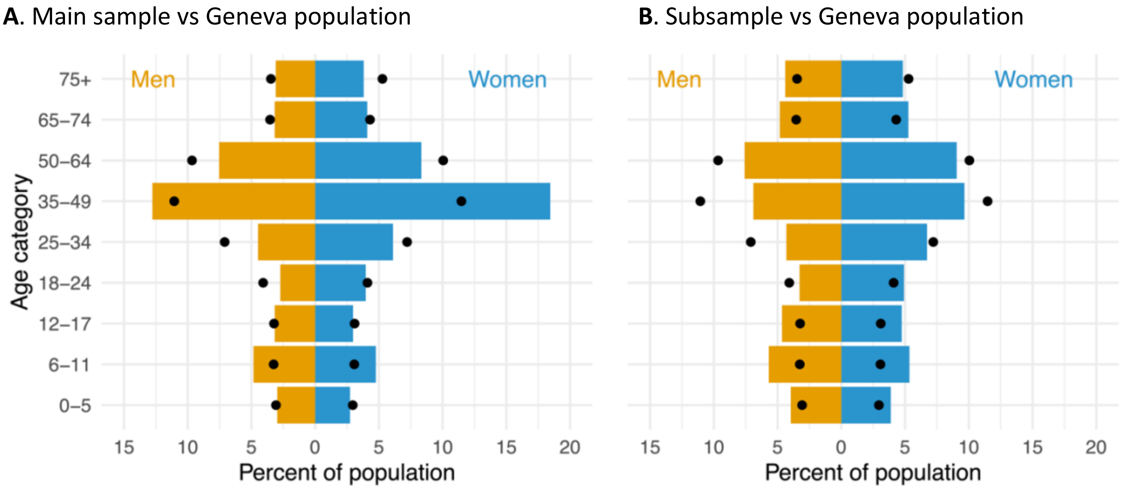


Dark yellow represents men; blue represents women. **A.** Total sample N =2521 used in estimations of seroprevalence of antibodies of vaccination and/or infection and infection-only origin; **B.** Subsample N =1160 used in estimation of seroprevalence of neutralizing antibodies.

**Supplementary figure 5.** Antibodies' response category and vaccination status

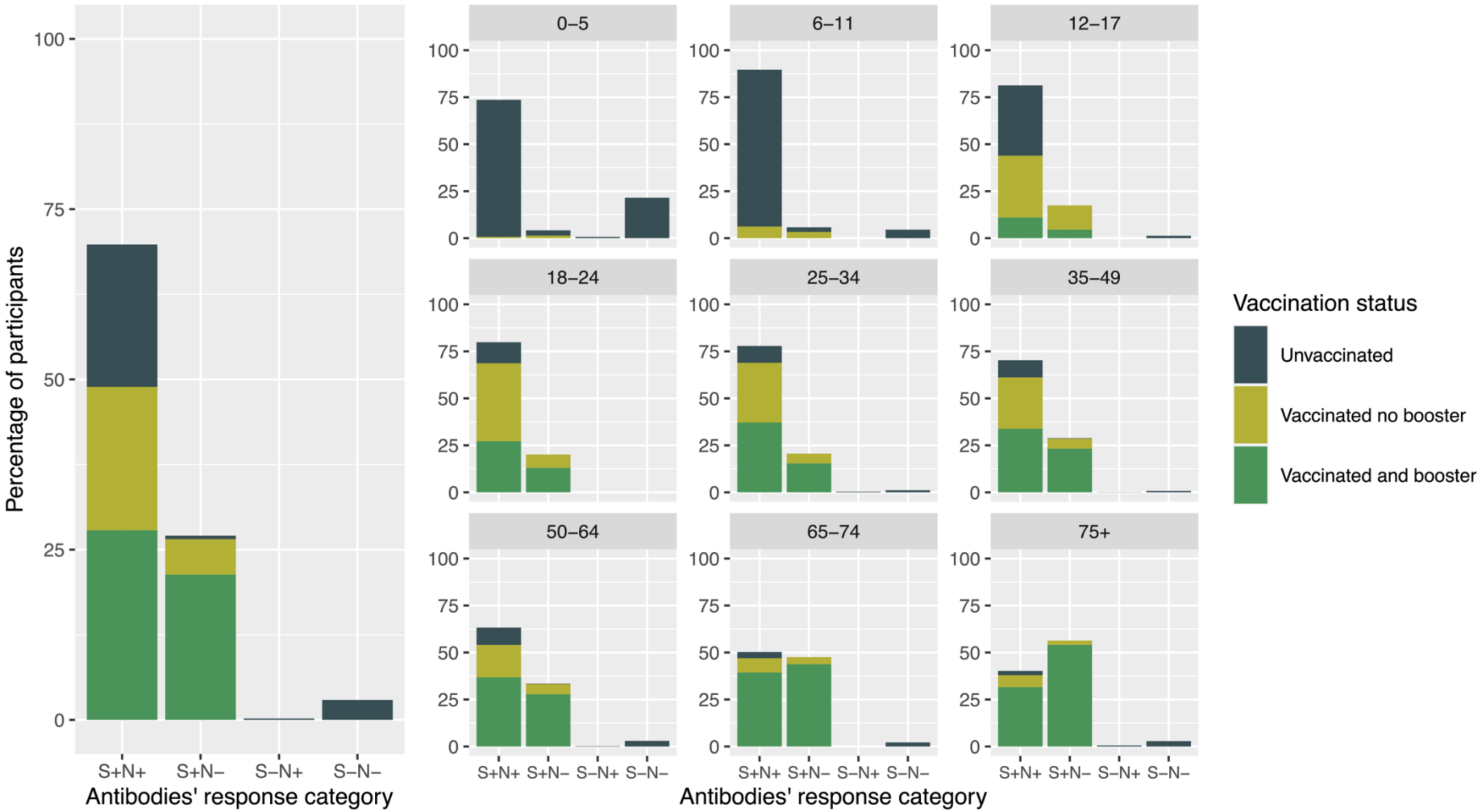


Number of participants in the four possible categories of the anti-S and anti-N serological tests. **+** indicates antibodies detected above positivity threshold; **-** indicates no antibodies detected above positivity threshold. Data on vaccination status was self-reported by the participants at the moment of the blood drawing.

**Supplementary figure 6.** Confirmed SARS-CoV-2 infection cases and estimated seroprevalence of anti-SARS-CoV-2 antibodies in the general population of Geneva, Switzerland, April 29th to June 9th, 2022

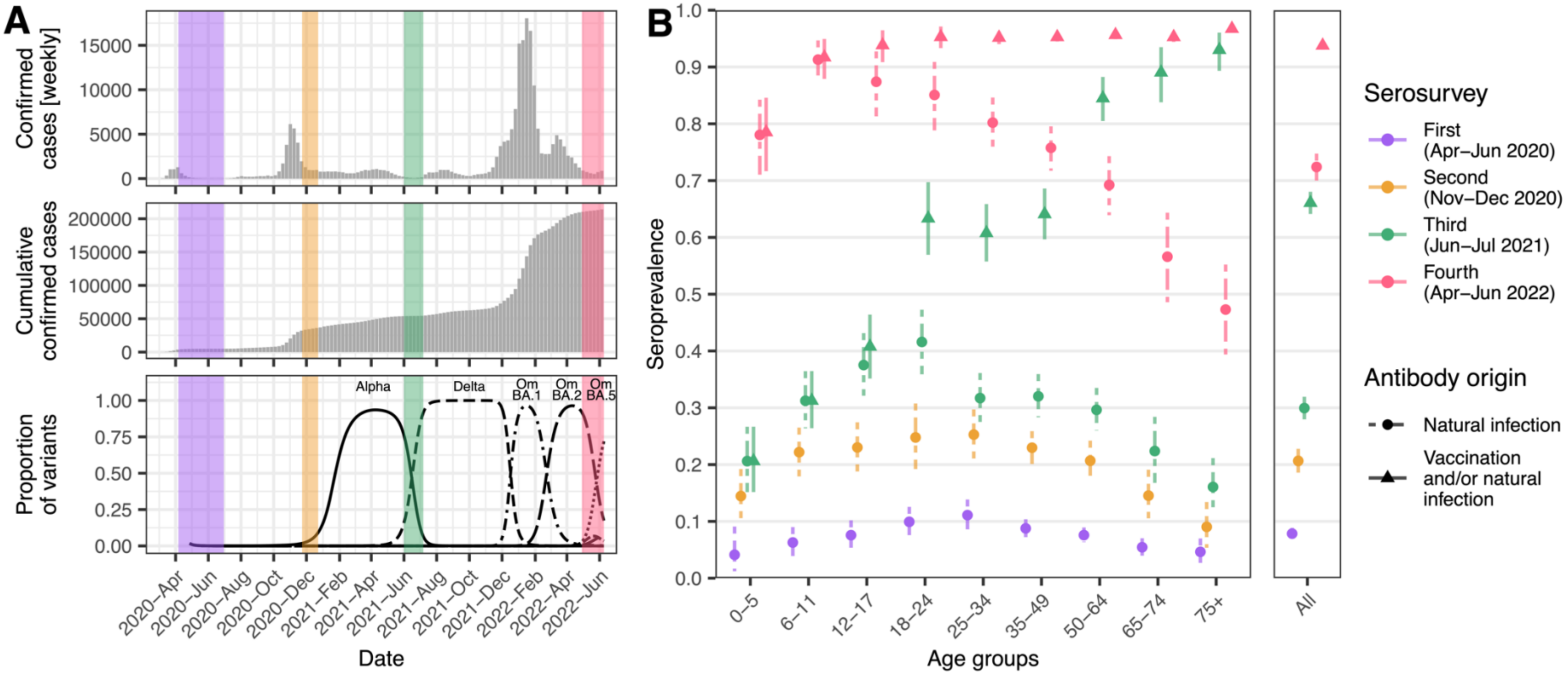


**A. Confirmed SARS-CoV-2 cases in the general population of Geneva**. Cantonal data presented in grey (available from: https://infocovid.smc.unige.ch) and serosurvey timings (colour shaded). Proportion of SARS-CoV-2 variants was estimated through multinomial spline regression of publicly available weekly sequence data from the Covariants project (https://github.com/hodcroftlab/covariants/), following analysis from https://www.hug.ch/laboratoire-virologie/surveillance-variants-sars-cov-2-geneve-national (report May 2022).
**B. Seroprevalence estimates by age group, serosurvey period and origin of antibody response**. Symbols indicate the antibody origin: *dot* indicates antibodies developed after infection (during the first two serosurveys, which took place before vaccination became available, infection was the only possible antibody origin). For first serosurvey the 0-5 group only included children aged 5 years (5); *triangle* indicates antibodies developed after infection and/or vaccination. Vertical bars represent 95% credible intervals in seroprevalence estimations.

**Supplementary figure 7.** Distribution of ED50% values for all tested SARS-CoV-2 variants by age group

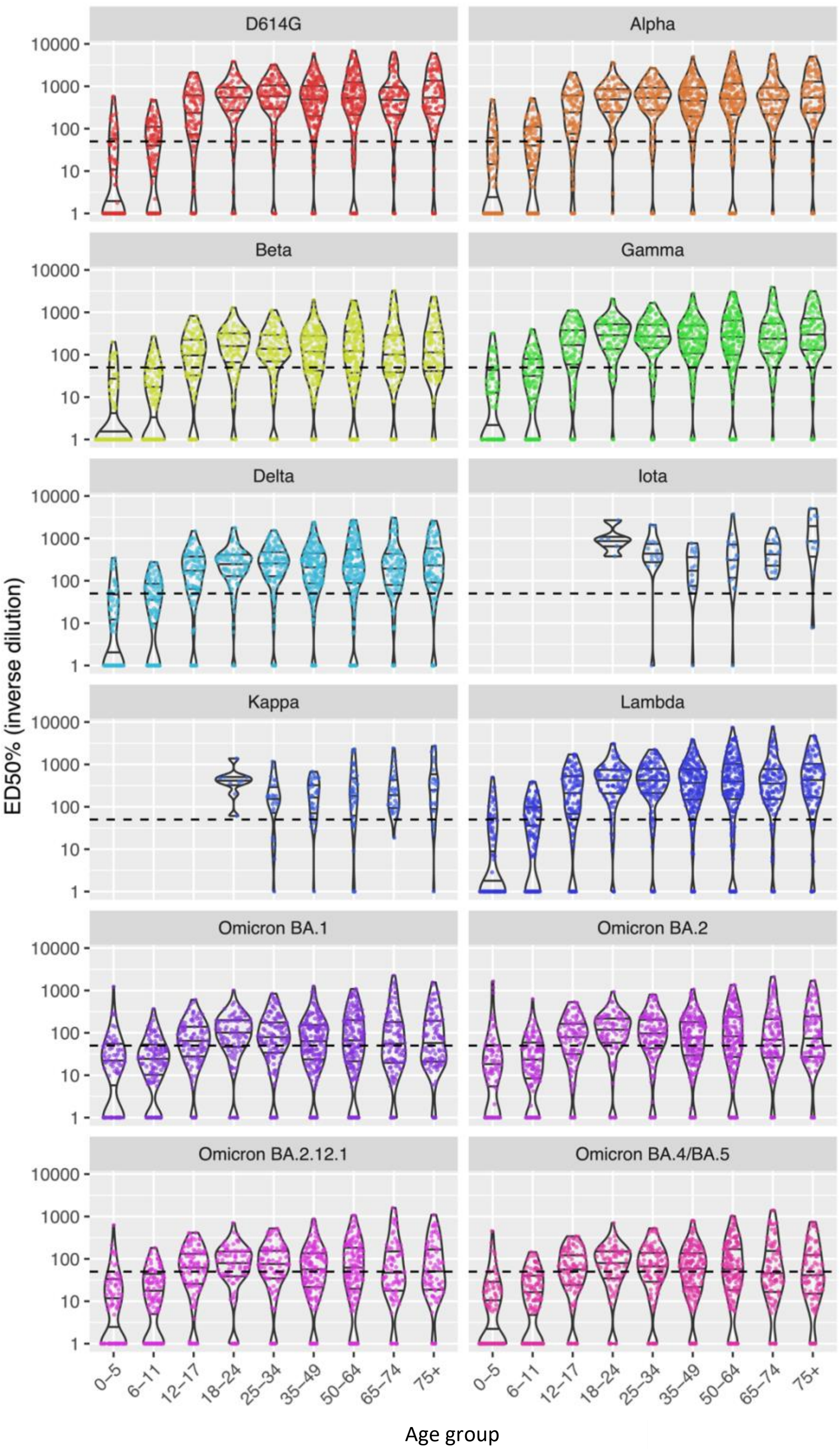


ED50%: mean serum dilution needed to achieve 50% neutralization.

The cell-free surrogate neutralization assay can be multiplexed to test up to ten variants on the same sample. For logistics reasons, numbers of tested samples per variant were as follows: N=1160 for D614G, Alpha, Beta, Gamma, Delta, Lambda, Omicron BA.1, and Omicron BA.2; N=1078 for Omicron BA.4/BA.5; N=995 for Omicron BA.2.12.1; N=165 for Kappa; and N=82 for Iota.

Black horizontal bars in each violin indicate quartiles.

**Supplementary figure 8.** Seroprevalence of neutralizing antibodies against main SARS-CoV-2 variants in the general population of Geneva, Switzerland, April 29th to June 9th, 2022

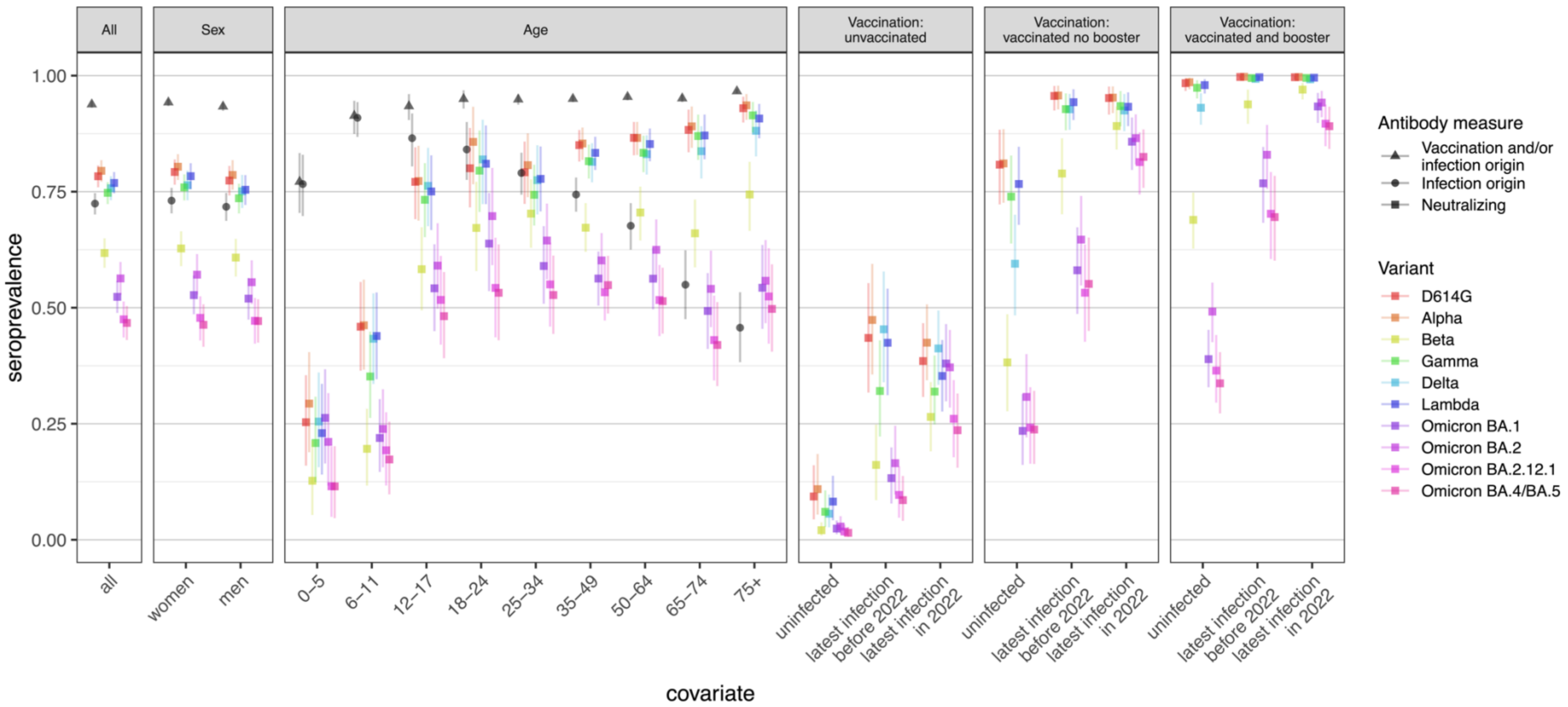


Panels show the effect of covariates tested in the model: sex, age, and infection and vaccination statuses (self-reported).
Global seroprevalence estimates for anti-S and anti-N antibodies are included in black in the three left panels for comparison purposes. Symbols indicate the antibody origin: *dot* indicates antibodies developed after infection (anti-N); *triangle* indicates antibodies developed after infection and/or vaccination (anti-S); and *square* indicates neutralizing antibodies (cell-free surrogate neutralization assay). Vertical bars represent 95% credible intervals.

**Supplementary table 1.** Comparison of education level in the main study sample and in Geneva population

| Education level | Study sample N (%) | Geneva population N (%) |
|---|---|---|
| Primary | 213 (11.3) | 98246 (26.6) |
| Secondary | 597 (31.7) | 118125 (32.0) |
| Tertiary | 1071 (56.9) | 153334 (41.5) |

Primary education corresponds to mandatory schooling; secondary education includes high-school diploma or apprenticeship training; tertiary education includes university-level diploma. Geneva population data available from: https://www.ge.ch/statistique/domaines/15/15_03/tableaux.asp#1

**Supplementary table 2.** Comparison of proportion vaccinated in main study sample and in Geneva population

| | Study sample | | Geneva population | |
|---|---|---|---|---|
| | **N** | **Vaccinated (self-reported) N (%)** | **N** | **Vaccinated N (%)** |
| **Total** | 2521 | 1902 (75.4) | 506343 | 360010 (71.1) |
| **Men** | 1129 | 844 (74.8) | 245320 | 174423 (69.6) |
| **Women** | 1392 | 1058 (76.0) | 261023 | 185587 (72.5) |
| **Age, y** | | | | |
| 0-9 | 304 | 18 (5.9) | 53083 | 1152 (2.2) |
| 10-19 | 280 | 150 (53.6) | 53405 | 27803 (52.1) |
| 20-29 | 219 | 198 (90.4) | 64680 | 47779 (73.9) |
| 30-39 | 399 | 360 (90.2) | 76299 | 58788 (77.1) |
| 40-49 | 547 | 485 (88.7) | 76578 | 62985 (82.3) |
| 50-59 | 278 | 242 (87.1) | 72233 | 61347 (84.9) |
| 60-69 | 212 | 190 (89.6) | 47544 | 42181 (88.7) |
| 70-79 | 181 | 175 (96.7) | 36849 | 33054 (89.7) |
| ≥80 | 71 | 64 (90.1) | 25672 | 24848 (96.8) |

Data on individuals vaccinated with at least 1 dose in study sample to match data on individuals vaccinated in the general population of Geneva with at least 1 dose up to May 8$^{th}$, 2022.
Geneva population data available from:
https://www.covid19.admin.ch/en/vaccination/persons/d/demography?geo=GE&demoSum=total&demoAge=minOne

**Supplementary table 3.** Prevalence ratio for seroprevalence of anti-SARS-CoV-2 antibodies in Geneva, Switzerland, April 29th to June 9th, 2022

| | | **Antibodies of any origin** | | **Antibodies of infection origin** | |
|---|---|---|---|---|---|
| | **Participants n (%)** | **Prevalence ratio (95% CrI)[a]** | ***P value*** | **Prevalence ratio (95% CrI)[a]** | ***P value*** |
| **Men** | 1129 (44.8) | 0.99 (0.98-1.00) | *0.13* | 0.97 (0.93-1.01) | *0.13* |
| **Women** | 1392 (55.2) | 1.00 (ref.) | | 1.00 (ref.) | |
| **Age**, y | | | | | |
| 0-5 | 144 (5.7) | 0.83 (0.75-0.89) | *< 0.001* | 0.97 (0.88-1.07) | *0.57* |
| 6-11 | 242 (9.6) | 0.96 (0.92-1.00) | *0.03* | 1.14 (1.07-1.21) | *< 0.0001* |
| 12-17 | 155 (6.2) | 0.99 (0.95-1.01) | *0.36* | 1.09 (1.00-1.18) | *0.04* |
| 18-24 | 169 (6.7) | 1.00 (0.98-1.02) | *0.90* | 1.06 (0.98-1.15) | *0.17* |
| 25-34 | 267 (10.6) | 1.00 (ref.) | | 1.00 (ref.) | |
| 35-49 | 787 (31.2) | 1.00 (0.99-1.01) | *0.86* | 0.95 (0.88-1.01) | *0.10* |
| 50-64 | 400 (15.9) | 1.01 (0.99-1.02) | *0.44* | 0.86 (0.79-0.94) | *< 0.001* |
| 65-74 | 183 (7.3) | 1.00 (0.99-1.02) | *0.87* | 0.71 (0.61-0.80) | *< 0.0001* |
| ≥75 | 174 (6.9) | 1.02 (1.01-1.03) | *<0.01* | 0.59 (0.49-0.69) | *< 0.0001* |
| **Education level[b]** | | | | | |
| Primary | 213 (11.3) | 1.01 (0.99-1.02) | *0.23* | 1.05 (0.97-1.13) | *0.28* |
| Secondary | 597 (31.7) | 1.01 (1.00-1.02) | *0.21* | 1.04 (0.98-1.09) | *0.24* |
| Tertiary | 1071 (56.9) | 1.00 (ref.) | | 1.00 (ref.) | |

[a] Prevalence ratio (95% credible interval) from Bayesian multinomial regression models accounting for sex, age, test performance and household clustering. Reference group for age and sex estimates (N = 2521) is female, aged 24-35 years. For education level estimates (N =1881), reference group is female, aged 25-44 years with tertiary education level.

[b] Self-reported education level among participants aged ≥18 years. Primary education corresponds to mandatory schooling; secondary education includes high-school diploma or apprenticeship training; tertiary education includes university-level diploma.

**Supplementary table 4.** Educational level of sample, serological results, and seroprevalence estimates in Geneva, Switzerland, April 29th to June 9th, 2022

| | | | Seropositive[b] | | Seroprevalence[c] | |
|---|---|---|---|---|---|---|
| | Participants | Vaccinated (self-reported)[a] | Anti-SARS-CoV-2 S protein | Anti-SARS-CoV-2 N protein | Antibodies of any origin | Antibodies of infection origin |
| | n (%) | n (%) | n (%) | n (%) | % (95% CrI) | % (95% CrI) |
| **Total** | 1881 | 1699 (90.3) | 1849 (98.3) | 1239 (65.9) | 95.6 (95.0-96.2) | 71.8 (68.5-75.1) |
| **Education[d]** | | | | | | |
| Primary | 213 (11.3) | 192 (90.1) | 209 (98.1) | 142 (66.7) | 96.0 (94.8-97.1) | 73.1 (66.2-80.0) |
| Secondary | 597 (31.7) | 508 (85.1) | 578 (96.8) | 402 (67.3) | 95.8 (95.0-96.6) | 72.4 (68.1-76.8) |
| Tertiary | 1071 (56.9) | 999 (93.3) | 1062 (99.2) | 695 (64.9) | 95.2 (94.5-95.9) | 70.4 (66.8-74.1) |

CrI: credible interval; N: nucleocapsid protein; S: spike protein; SARS-CoV-2: severe acute respiratory syndrome coronavirus 2.

[a] Self-reported having received at least 1 dose of the COVID-19 vaccine before blood sample was drawn.

[b] Serology based on Roche Elecsys anti-SARS-CoV-2 S immunoassay and N immunoassay, respectively.

[c] Seroprevalence estimates reported as % and 95% credible interval, adjusted for test performance of both immunoassays and post-stratified to account for age distribution in the Geneva general population and for household clustering of infection and vaccination. Seroprevalence of antibodies of any antibodies is based on proportion of participants with any anti-SARS-CoV-2 antibodies, additionally post-stratified to the vaccination data in the general population of Geneva; seroprevalence of infection antibodies is based on proportion of participants who were naturally infected (but could also have been vaccinated).

[d] Self-reported education level among participants aged ≥18 years. Primary education corresponds to mandatory schooling; secondary education includes high-school diploma or apprenticeship training; tertiary education includes university-level diploma.

**Supplementary table 5.** Characteristics of main sample and subsample analyzed in this study

| | **Total seroprevalence sample** | **Neutralizing antibodies sample** |
|---|---|---|
| **Total** | 2521 | 1160 |
| **Men** | 1129 (44.8) | 528 (45.6) |
| **Women** | 1392 (55.2) | 632 (54.4) |
| **Age**, y | | |
| 0-5 | 144 (5.7) | 91 (7.9) |
| 6-11 | 242 (9.6) | 128 (11.0) |
| 12-17 | 155 (6.2) | 109 (9.4) |
| 18-24 | 169 (6.7) | 95 (8.2) |
| 25-34 | 267 (10.6) | 128 (11.0) |
| 35-49 | 787 (31.2) | 192 (16.6) |
| 50-64 | 400 (15.9) | 193 (16.7) |
| 65-74 | 183 (7.3) | 117 (10.1) |
| ≥75 | 174 (6.9) | 107 (9.2) |
| **Seropositive for anti-N antibodies** | 1765 (70.0) | 811 (70.0) |

**Supplementary table 6.** Demographic characteristics of subsample, serological results, and seroprevalence estimates in Geneva, Switzerland, April 29th to June 9th, 2022

| | | | **Seropositive[b]** | | **Seroprevalence[c]** | |
|---|---|---|---|---|---|---|
| | **Participants** | **Vaccinated (self-reported)[a]** | **Anti-SARS-CoV-2 S protein** | **Anti-SARS-CoV-2 N protein** | **Antibodies of any origin** | **Antibodies of infection origin** |
| | n (%) | n (%) | n (%) | n (%) | % (95% CrI) | % (95% CrI) |
| **Total** | 1160 | 831 (71.6) | 1119 (96.5) | 813 (70.1) | 94.4 (93.5-95.3) | 74.3 (70.9-77.7) |
| **Men** | 528 (45.6) | 372 (70.3) | 506 (95.7) | 359 (67.9) | 93.8 (92.6-95.0) | 72.8 (68.2-77.4) |
| **Women** | 632 (54.4) | 459 (72.7) | 613 (97.1) | 454 (71.9) | 95.0 (94.0-96.0) | 75.7 (71.6-79.8) |
| **Age**, y | | | | | | |
| 0-5 | 91 (7.9) | 3 (3.3) | 72 (79.1) | 67 (73.6) | 78.2 (69.4-85.9) | 77.7 (68.7-85.6) |
| 6-11 | 128 (11.0) | 13 (10.2) | 124 (96.9) | 115 (89.8) | 94.5 (90.0-97.9) | 94.2 (89.5-97.8) |
| 12-17 | 109 (9.4) | 69 (63.3) | 107 (98.2) | 90 (82.6) | 94.2 (90.4-97.4) | 88.2 (80.4-94.7) |
| 18-24 | 95 (8.2) | 85 (89.5) | 95 (100.0) | 79 (83.2) | 96.6 (93.9-98.8) | 89.3 (80.7-96.1) |
| 25-34 | 128 (11.0) | 118 (92.2) | 128 (100.0) | 103 (80.5) | 95.7 (94.1-97.1) | 82.6 (76.0-88.3) |
| 35-49 | 192 (16.6) | 168 (87.5) | 191 (99.5) | 133 (69.3) | 95.2 (93.7-96.6) | 75.2 (68.0-82.5) |
| 50-64 | 193 (16.7) | 165 (85.5) | 183 (94.8) | 121 (62.7) | 95.4 (94.3-96.5) | 67.7 (60.0-75.2) |
| 65-74 | 117 (10.1) | 110 (94.0) | 115 (98.3) | 58 (49.6) | 95.1 (94.0-96.2) | 55.3 (45.2-65.1) |
| ≥75 | 107 (9.2) | 100 (93.5) | 104 (97.2) | 47 (43.9) | 96.9 (96.3-97.5) | 49.7 (39.9-60.0) |

CrI: credible interval; N: nucleocapsid protein; S: spike protein; SARS-CoV-2: severe acute respiratory syndrome coronavirus 2.

[a] Self-reported having received at least 1 dose of the COVID-19 vaccine before blood sample was drawn.

[b] Serology based on Roche Elecsys anti-SARS-CoV-2 S immunoassay and N immunoassay, respectively.

[c] Seroprevalence estimates reported as % and 95% credible interval, adjusted for test performance of both immunoassays and post-stratified to account for age distribution in the Geneva general population and for household clustering of infection and vaccination. Seroprevalence of antibodies of any origin is based on proportion of participants with any anti-SARS-CoV-2 antibodies, additionally post-stratified to the vaccination data in the general population of Geneva; seroprevalence of infection antibodies is based on proportion of participants who were naturally infected (but could have also been vaccinated).

**Supplementary table 7.** Seroprevalence estimates of neutralizing anti-SARS-CoV-2 antibodies against main variants in the general population of Geneva, Switzerland, April 29th to June 9th, 2022

| | | SARS-CoV-2 variant | | | | | | | |
|---|---|---|---|---|---|---|---|---|---|
| | | D614G | | Alpha | | Beta | | Gamma | |
| | n | n (%)[a] | Seroprevalence % (95% CrI)[b] | n (%)[a] | Seroprevalence % (95% CrI)[b] | n (%)[a] | Seroprevalence % (95% CrI)[b] | n (%)[a] | Seroprevalence % (95% CrI)[b] |
| **Total** | 1160 | 910 (78.4) | 78.3 (76.0-80.7) | 918 (79.1) | 79.5 (77.1-81.8) | 704 (60.7) | 61.8 (58.6-64.9) | 871 (75.1) | 74.8 (72.3-77.1) |
| **Men** | 631 | 506 (80.2) | 79.2 (76.5-81.9) | 510 (80.8) | 80.4 (77.5-83.1) | 395 (62.6) | 62.8 (59.0-66.5) | 485 (76.9) | 75.9 (73.1-78.7) |
| **Women** | 529 | 404 (76.4) | 77.4 (74.2-80.6) | 408 (77.1) | 78.6 (75.4-81.8) | 309 (58.4) | 60.8 (56.7-64.8) | 386 (73.0) | 73.6 (70.3-76.7) |
| **Age group**, y | | | | | | | | | |
| 0-5 | 91 | 19 (20.9) | 25.3 (16.0-35.5) | 22 (24.2) | 29.3 (18.9-40.4) | 10 (11.0) | 12.7 (5.3-21.4) | 16 (17.6) | 20.9 (12.4-30.8) |
| 6-11 | 128 | 55 (43.0) | 45.9 (36.4-55.5) | 55 (43.0) | 46.2 (36.7-56.0) | 25 (19.5) | 19.6 (11.7-28.2) | 43 (33.6) | 35.2 (26.3-44.8) |
| 12-17 | 109 | 88 (80.7) | 77.1 (69.1-84.6) | 88 (80.7) | 77.3 (68.8-84.9) | 70 (64.2) | 58.3 (49.4-67.3) | 85 (78.0) | 73.2 (65.2-81.1) |
| 18-24 | 95 | 86 (90.5) | 80.0 (71.6-88.7) | 89 (93.7) | 85.7 (76.6-93.3) | 75 (78.9) | 67.2 (57.9-77.1) | 85 (89.5) | 79.5 (70.6-88.2) |
| 25-34 | 128 | 116 (90.6) | 79.2 (72.4-85.8) | 117 (91.4) | 80.7 (73.8-87.6) | 103 (80.5) | 70.3 (62.9-77.7) | 112 (87.5) | 74.3 (67.7-80.8) |
| 35-49 | 192 | 176 (91.7) | 85.0 (81.5-88.3) | 175 (91.1) | 85.4 (81.9-88.8) | 134 (69.8) | 67.2 (62.0-72.5) | 169 (88.0) | 81.6 (77.8-85.1) |
| 50-64 | 193 | 165 (85.5) | 86.6 (82.8-90.0) | 165 (85.5) | 86.6 (82.9-90.0) | 131 (67.9) | 70.5 (64.5-76.1) | 159 (82.4) | 83.4 (79.2-87.0) |
| 65-74 | 117 | 106 (90.6) | 88.3 (83.5-92.7) | 107 (91.5) | 89.1 (84.5-93.3) | 79 (67.5) | 66.0 (58.7-73.3) | 105 (89.7) | 87.0 (81.9-91.6) |
| ≥75 | 107 | 99 (92.5) | 93.0 (89.9-95.5) | 100 (93.5) | 93.6 (90.5-96.0) | 77 (72.0) | 74.4 (66.6-81.4) | 97 (90.7) | 91.4 (87.7-94.3) |
| **Vaccination and infection status**[c] | | | | | | | | | |
| Unvaccinated and uninfected | 39 | 0 (0.0) | 9.3 (4.4-16.1) | 0 (0.0) | 10.9 (5.4-18.5) | 0 (0.0) | 2.1 (0.9-3.8) | 0 (0.0) | 6.0 (2.8-10.6) |
| Unvaccinated and pre-2022 infection | 55 | 29 (52.7) | 43.5 (31.7-55.3) | 30 (54.5) | 47.4 (35.6-59.4) | 16 (29.1) | 16.1 (8.5-24.8) | 22 (40.0) | 32.1 (22.3-43.0) |
| Unvaccinated and 2022 infection | 94 | 27 (28.7) | 38.5 (30.8-46.7) | 29 (30.9) | 42.5 (34.4-50.7) | 16 (17.0) | 26.5 (19.1-34.0) | 23 (24.5) | 31.9 (24.9-39.7) |
| Vaccinated, no booster, and uninfected | 54 | 38 (70.4) | 80.8 (72.3-88.4) | 38 (70.4) | 81.1 (72.6-88.5) | 11 (20.4) | 38.2 (27.7-48.6) | 34 (63.0) | 73.9 (63.8-82.8) |
| Vaccinated, no booster, and pre-2022 infection | 77 | 76 (98.7) | 95.6 (92.5-97.8) | 76 (98.7) | 95.7 (92.7-97.8) | 60 (77.9) | 78.9 (70.1-86.5) | 73 (94.8) | 92.8 (88.2-96.2) |
| Vaccinated, no booster, and 2022 infection | 93 | 93 (100.0) | 95.2 (91.6-97.7) | 93 (100.0) | 95.3 (91.9-97.7) | 86 (92.5) | 89.1 (84.1-93.4) | 91 (97.8) | 93.4 (89.1-96.7) |
| Vaccinated, booster, and uninfected | 224 | 221 (98.7) | 98.4 (96.7-99.4) | 222 (99.1) | 98.6 (97.1-99.5) | 140 (62.5) | 68.9 (62.7-74.8) | 216 (96.4) | 97.4 (95.1-99.0) |
| Vaccinated, booster, and pre-2022 infection | 73 | 73 (100.0) | 99.7 (99.3-99.9) | 73 (100.0) | 99.7 (99.4-99.9) | 62 (84.9) | 93.8 (89.6-97.0) | 72 (98.6) | 99.5 (98.8-99.8) |
| Vaccinated, booster, and 2022 infection | 104 | 103 (99.0) | 99.7 (99.2-99.9) | 103 (99.0) | 99.7 (99.3-99.9) | 101 (97.1) | 97.0 (94.9-98.4) | 103 (99.0) | 99.5 (98.9-99.9) |

[a] Number and proportion (%) above the threshold for neutralizing activity against specific variant. [b] Estimated seroprevalence % (95% credible interval) of neutralizing antibodies against specific variant, from regression model accounting for age, sex, self-reported most recent SARS-CoV-2 infection and vaccination status, and post-stratifying to the demographic and vaccination and infection statuses of the Geneva general population. [c] Self-reported vaccination and most recent infection via a COVID-19 diagnostic positive test (RT-PCR or rapid antigen test, including self-test). The cell-free surrogate neutralization assay can be multiplexed to test up to ten variants on the same sample. For logistics reasons, numbers of tested samples per variant were as follows: N=1160 for D614G, Alpha, Beta, Gamma, Delta, Lambda, Omicron BA.1, and Omicron BA.2; N=1078 for Omicron BA.4/BA.5; N=995 for Omicron BA.2.12.1; N=165 for Kappa; and N=82 for Iota.

**Supplementary table 7 continued.** Seroprevalence estimates of neutralizing anti-SARS-CoV-2 antibodies against main variants in the general population of Geneva, Switzerland, April 29th to June 9th, 2022

| | | SARS-CoV-2 variant | | | | | | | |
|---|---|---|---|---|---|---|---|---|---|
| | | Delta | | Iota | | Kappa | | Lambda | |
| | n | n (%)[a] | Seroprevalence % (95% CrI)[b] | n (%)[a] | Seroprevalence % (95% CrI)[d] | n (%)[a] | Seroprevalence % (95% CrI)[d] | n (%)[a] | Seroprevalence % (95% CrI)[b] |
| **Total** | 1160 | 862 (74.3) | 75.7 (73.2-78.3) | 78 (95.1) | - | 140 (84.8) | - | 893 (77.0) | 76.9 (74.6-79.2) |
| **Men** | | 477 (75.6) | 76.4 (73.2-79.5) | 43 (95.6) | - | 81 (85.3) | - | 499 (79.1) | 78.3 (75.5-81.1) |
| **Women** | | 385 (72.8) | 75.1 (71.5-78.6) | 35 (94.6) | - | 59 (84.3) | - | 394 (74.5) | 75.4 (72.1-78.6) |
| **Age group**, y | | | | | | | | | |
| 0-5 | 91 | 18 (19.8) | 25.5 (15.6-36.0) | - | - | - | - | 17 (18.7) | 23.0 (14.1-33.6) |
| 6-11 | 128 | 49 (38.3) | 43.3 (33.6-53.1) | - | - | - | - | 52 (40.6) | 43.9 (34.6-53.3) |
| 12-17 | 109 | 85 (78.0) | 76.3 (67.6-84.4) | - | - | - | - | 86 (78.9) | 75.0 (66.7-82.8) |
| 18-24 | 95 | 85 (89.5) | 82.0 (72.3-90.5) | 5 (100.0) | - | 9 (100.0) | - | 86 (90.5) | 81.0 (72.3-89.3) |
| 25-34 | 128 | 112 (87.5) | 77.5 (70.1-85.0) | 14 (93.3) | - | 20 (80.0) | - | 115 (89.8) | 77.8 (70.8-84.7) |
| 35-49 | 192 | 165 (85.9) | 81.4 (77.0-85.4) | 15 (93.8) | - | 29 (85.3) | - | 173 (90.1) | 83.4 (79.7-86.8) |
| 50-64 | 193 | 156 (80.8) | 83.1 (78.7-87.1) | 16 (94.1) | - | 32 (76.2) | - | 163 (84.5) | 85.3 (81.6-88.6) |
| 65-74 | 117 | 100 (85.5) | 83.7 (77.9-89.1) | 18 (100.0) | - | 29 (96.7) | - | 105 (89.7) | 87.1 (82.0-91.6) |
| ≥75 | 107 | 92 (86.0) | 88.1 (82.6-92.5) | 10 (90.9) | - | 21 (84.0) | - | 96 (89.7) | 90.8 (86.5-93.9) |
| **Vaccination and infection status**[c] | | | | | | | | | |
| Unvaccinated and uninfected | 39 | 0 (0.0) | 5.6 (2.7-9.7) | 0 (0.0) | - | 0 (0.0) | - | 0 (0.0) | 8.2 (4.2-13.8) |
| Unvaccinated and pre-2022 infection | 55 | 29 (52.7) | 45.3 (34.0-57.8) | - | - | 0 (0.0) | - | 29 (52.7) | 42.4 (31.2-54.1) |
| Unvaccinated and 2022 infection | 94 | 30 (31.9) | 41.2 (33.4-49.4) | 3 (100.0) | - | 1 (25.0) | - | 25 (26.6) | 35.3 (27.6-43.0) |
| Vaccinated, no booster, and uninfected | 54 | 22 (40.7) | 59.5 (48.4-70.0) | 1 (50.0) | - | 2 (28.6) | - | 35 (64.8) | 76.7 (67.9-84.7) |
| Vaccinated, no booster, and pre-2022 infection | 77 | 73 (94.8) | 92.8 (88.3-96.1) | 6 (100.0) | - | 10 (90.9) | - | 74 (96.1) | 94.3 (90.4-97.1) |
| Vaccinated, no booster, and 2022 infection | 93 | 88 (94.6) | 92.5 (88.0-96.0) | 4 (100.0) | - | 9 (100.0) | - | 91 (97.8) | 93.3 (89.1-96.4) |
| Vaccinated, booster, and uninfected | 224 | 203 (90.6) | 93.1 (89.4-96.2) | 23 (100.0) | - | 45 (93.8) | - | 219 (97.8) | 98.0 (96.1-99.3) |
| Vaccinated, booster, and pre-2022 infection | 73 | 72 (98.6) | 99.3 (98.6-99.7) | 6 (100.0) | - | 12 (100.0) | - | 73 (100.0) | 99.7 (99.2-99.9) |
| Vaccinated, booster, and 2022 infection | 104 | 102 (98.1) | 99.2 (98.4-99.7) | 13 (100.0) | - | 26 (100.0) | - | 103 (99.0) | 99.6 (99.0-99.9) |

[a] Number and proportion (%) above the threshold for neutralizing activity against specific variant. [b] Estimated seroprevalence % (95% credible interval) of neutralizing antibodies against specific variant, from regression model accounting for age, sex, self-reported most recent SARS-CoV-2 infection and vaccination status, and post-stratifying to the demographic and vaccination and infection statuses of the Geneva general population. [c] Self-reported vaccination and most recent infection via a COVID-19 diagnostic positive test (RT-PCR or rapid antigen test, including self-test). The cell-free surrogate neutralization assay can be multiplexed to test up to ten variants on the same sample. For logistics reasons, numbers of tested samples per variant were as follows: N=1160 for D614G, Alpha, Beta, Gamma, Delta, Lambda, Omicron BA.1, and Omicron BA.2; N=1078 for Omicron BA.4/BA.5; N=995 for Omicron BA.2.12.1; N=165 for Kappa; and N=82 for Iota.

**Supplementary table 7 continued.** Seroprevalence estimates of neutralizing anti-SARS-CoV-2 antibodies against main variants in the general population of Geneva, Switzerland, April 29th to June 9th, 2022

| | | SARS-CoV-2 variant | | | | | | | |
|---|---|---|---|---|---|---|---|---|---|
| | | Omicron BA.1 | | Omicron BA.2 | | Omicron BA.2.12.1 | | Omicron BA.4/BA.5 | |
| | n | n (%)[a] | Seroprevalence % (95% CrI)[b] | n (%)[a] | Seroprevalence % (95% CrI)[b] | n (%)[a] | Seroprevalence % (95% CrI)[b] | n (%)[a] | Seroprevalence % (95% CrI)[b] |
| **Total** | 1160 | 587 (50.6) | 52.3 (48.9-55.8) | 640 (55.2) | 56.3 (52.8-59.8) | 455 (45.7) | 47.5 (43.6-51.3) | 486 (45.1) | 46.7 (43.0-50.4) |
| **Men** | | 327 (51.8) | 52.7 (48.6-56.9) | 358 (56.7) | 57.1 (52.8-61.6) | 251 (46.8) | 47.8 (42.9-52.5) | 269 (45.9) | 46.3 (41.6-50.7) |
| **Women** | | 260 (49.1) | 52.0 (47.4-56.4) | 282 (53.3) | 55.5 (50.8-60.2) | 204 (44.4) | 47.2 (42.3-52.1) | 217 (44.1) | 47.1 (42.5-51.8) |
| **Age group**, y | | | | | | | | | |
| 0-5 | 91 | 19 (20.9) | 26.3 (16.4-36.7) | 17 (18.7) | 21.1 (11.7-31.6) | 9 (9.9) | 11.5 (5.0-20.1) | 9 (9.9) | 11.5 (4.7-20.2) |
| 6-11 | 128 | 26 (20.3) | 21.9 (14.6-30.3) | 30 (23.4) | 23.9 (15.6-32.4) | 25 (19.5) | 19.3 (11.7-27.5) | 23 (18.0) | 17.3 (9.8-25.5) |
| 12-17 | 109 | 65 (59.6) | 54.2 (45.0-63.7) | 70 (64.2) | 59.1 (49.8-68.2) | 63 (57.8) | 51.7 (42.0-61.1) | 60 (55.0) | 48.2 (39.1-57.7) |
| 18-24 | 95 | 69 (72.6) | 63.8 (53.6-74.7) | 75 (78.9) | 69.7 (59.2-80.1) | 55 (64.0) | 54.3 (43.6-65.1) | 57 (63.3) | 53.2 (43.0-63.6) |
| 25-34 | 128 | 84 (65.6) | 59.0 (50.8-67.6) | 93 (72.7) | 64.5 (56.1-72.4) | 65 (63.1) | 55.0 (45.9-64.0) | 67 (59.3) | 52.7 (44.4-61.2) |
| 35-49 | 192 | 105 (54.7) | 56.3 (50.5-62.1) | 116 (60.4) | 60.2 (54.0-66.1) | 83 (52.5) | 53.3 (47.2-59.5) | 97 (55.1) | 54.9 (48.8-61.2) |
| 50-64 | 193 | 104 (53.9) | 56.3 (49.6-62.9) | 116 (60.1) | 62.5 (55.9-69.0) | 76 (50.3) | 51.7 (43.9-59.2) | 87 (49.4) | 51.4 (44.4-58.6) |
| 65-74 | 117 | 60 (51.3) | 49.3 (41.1-57.5) | 66 (56.4) | 54.1 (45.8-62.3) | 39 (44.8) | 43.0 (34.3-52.6) | 43 (43.4) | 42.0 (33.1-51.2) |
| ≥75 | 107 | 55 (51.4) | 54.3 (45.5-63.5) | 57 (53.3) | 55.8 (46.7-64.6) | 40 (48.8) | 52.4 (42.3-62.8) | 43 (44.8) | 49.7 (40.5-59.3) |
| **Vaccination and infection status**[c] | | | | | | | | | |
| Unvaccinated and uninfected | 39 | 0 (0.0) | 2.4 (1.2-4.2) | 1 (2.6) | 2.9 (1.3-5.1) | 0 (0.0) | 1.8 (0.8-3.3) | 0 (0.0) | 1.5 (0.6-2.8) |
| Unvaccinated and pre-2022 infection | 55 | 12 (21.8) | 13.3 (7.8-19.9) | 15 (27.3) | 16.5 (9.8-24.5) | 12 (22.6) | 9.7 (4.8-15.8) | 10 (18.2) | 8.5 (4.1-13.7) |
| Unvaccinated and 2022 infection | 94 | 29 (30.9) | 38.0 (29.8-46.5) | 24 (25.5) | 37.2 (28.5-45.2) | 18 (20.0) | 26.1 (17.8-34.4) | 17 (18.7) | 23.6 (15.5-31.5) |
| Vaccinated, no booster, and uninfected | 54 | 6 (11.1) | 23.5 (16.1-31.4) | 10 (18.5) | 30.8 (22.1-40.0) | 6 (12.8) | 24.2 (16.4-32.9) | 7 (13.5) | 23.8 (16.3-32.1) |
| Vaccinated, no booster, and pre-2022 infection | 77 | 44 (57.1) | 58.1 (48.7-67.3) | 48 (62.3) | 64.7 (54.8-74.1) | 34 (51.5) | 53.2 (42.7-63.6) | 38 (53.5) | 55.1 (45.0-65.1) |
| Vaccinated, no booster, and 2022 infection | 93 | 81 (87.1) | 85.8 (79.7-91.0) | 84 (90.3) | 86.5 (80.9-91.6) | 68 (81.0) | 81.4 (74.4-87.8) | 71 (79.8) | 82.5 (75.8-88.4) |
| Vaccinated, booster, and uninfected | 224 | 73 (32.6) | 38.9 (32.9-45.2) | 95 (42.4) | 49.2 (42.6-55.4) | 54 (30.7) | 36.5 (29.6-44.1) | 56 (27.9) | 33.7 (27.2-40.4) |
| Vaccinated, booster, and pre-2022 infection | 73 | 51 (69.9) | 76.8 (68.3-84.6) | 57 (78.1) | 83.0 (75.6-89.4) | 38 (62.3) | 70.2 (60.5-79.2) | 43 (64.2) | 69.5 (60.1-78.4) |
| Vaccinated, booster, and 2022 infection | 104 | 92 (88.5) | 93.4 (89.8-96.3) | 95 (91.3) | 94.2 (91.0-96.7) | 67 (85.9) | 89.6 (84.8-93.8) | 76 (83.5) | 89.1 (84.2-93.3) |

[a] Number and proportion (%) above the threshold for neutralizing activity against specific variant. [b] Estimated seroprevalence % (95% credible interval) of neutralizing antibodies against specific variant, from regression model accounting for age, sex, self-reported most recent SARS-CoV-2 infection and vaccination status, and post-stratifying to the demographic and vaccination and infection statuses of the Geneva general population. [c] Self-reported vaccination and most recent infection via a COVID-19 diagnostic positive test (RT-PCR or rapid antigen test, including self-test). The cell-free surrogate neutralization assay can be multiplexed to test up to ten variants on the same sample. For logistics reasons, numbers of tested samples per variant were as follows: N=1160 for D614G, Alpha, Beta, Gamma, Delta, Lambda, Omicron BA.1, and Omicron BA.2; N=1078 for Omicron BA.4/BA.5; N=995 for Omicron BA.2.12.1; N=165 for Kappa; and N=82 for Iota.

**Supplementary table 8.** Comparison of seroprevalence of anti-SARS-CoV-2 antibodies developed through infection by June-July 2021 and April-June 2022, Geneva, Switzerland

| | Geneva population N | Seroprevalence of antibodies after infection % | | Percent increase | Absolute increase % points | Absolute increase N |
|---|---|---|---|---|---|---|
| | | Jun-Jul 2021 | Apr-Jun 2022 | | | |
| **Total** | 511921 | 29.9 | 72.4 | 142% | 42.5 | 216229 |
| **Men** | 248170 | 30.4 | 71.7 | 136% | 41.3 | 101869 |
| **Women** | 263751 | 29.5 | 73.1 | 148% | 43.6 | 114284 |
| **Age, y** | | | | | | |
| 0-5 | 30840 | 20.8 | 76.7 | 268% | 55.8 | 17093 |
| 6-11 | 32382 | 31.4 | 90.5 | 189% | 59.5 | 19064 |
| 12-17 | 32377 | 37.7 | 86.4 | 129% | 48.8 | 15482 |
| 18-24 | 41899 | 41.8 | 84.0 | 101% | 42.3 | 17835 |
| 25-34 | 73273 | 31.9 | 78.9 | 148% | 47.1 | 34517 |
| 35-49 | 115343 | 32.2 | 74.4 | 131% | 42.2 | 48646 |
| 50-64 | 100883 | 29.8 | 67.7 | 127% | 37.9 | 37840 |
| 65-74 | 40143 | 22.5 | 55.0 | 144% | 32.5 | 13103 |
| ≥75 | 44781 | 16.2 | 45.9 | 182% | 29.5 | 12831 |

Percent increase calculated as: ((Apr-Jun 2022 seroprevalence / Jun-Jul 2021 seroprevalence) – 1) x 100.
Absolute increase calculated as: Apr-Jun 2022 seroprevalence – Jun-Jul 2021 seroprevalence.
Absolute increase N calculated as: absolute increase % x Geneva population
Seroprevalence estimates for Jun-Jul 2021 from previous seroprevalence study (1).
Data on Geneva population available from: https://www.ge.ch/statistique/domaines/01/01_01/tableaux.asp#5
